# Safety and immunogenicity of SARS-CoV-2 recombinant protein vaccine formulations in healthy adults: a randomised, placebo-controlled, dose-ranging study

**DOI:** 10.1101/2021.01.19.20248611

**Authors:** Paul A Goepfert, Bo Fu, Anne-Laure Chabanon, Matthew I Bonaparte, Matthew G Davis, Brandon J Essink, Ian Frank, Owen Haney, Helene Janosczyk, Michael C Keefer, Marguerite Koutsoukos, Murray A Kimmel, Roger Masotti, Stephen J Savarino, Lode Schuerman, Howard Schwartz, Lawrence D Sher, Jon Smith, Fernanda Tavares-Da-Silva, Sanjay Gurunathan, Carlos A DiazGranados, Guy De Bruyn

**Author notes:** **Corresponding Author:** Guy de Bruyn, Sanofi Pasteur, Swiftwater, PA, USA. **ClinicalTrials.gov:** NCT04537208.

## Abstract

**Background:** Effective vaccines against severe acute respiratory syndrome coronavirus 2 (SARS-CoV-2) are urgently needed. CoV2 preS dTM is a stabilised pre-fusion S protein vaccine produced in a baculovirus expression system. We present interim safety and immunogenicity results of the first-in-human study of the CoV2 preS dTM vaccine with two different adjuvant formulations.

**Methods:** This Phase I/II, randomised, double-blind study (NCT04537208) is being conducted in healthy, SARS-CoV-2-seronegative adults in the USA. Participants were stratified by age (18–49 and ≥50 years) and randomised to receive one (on Day[D]1) or two doses (D1, D22) of placebo or candidate vaccine, containing: low-dose (LD, effective dose 1.3 µg) or high-dose (HD, 2.6 µg) antigen with adjuvant AF03 (Sanofi Pasteur) or AS03 (GlaxoSmithKline); or unadjuvanted HD (18–49 years only). Safety was assessed up to D43. SARS-CoV-2 neutralising and binding antibody profiles were assessed in D1, D22 and D36 serum samples.

**Findings:** The interim safety analyses included 439/441 randomised participants. There were no related unsolicited immediate AEs, serious AEs, medically attended AEs classified as severe, or AE of special interest. More grade 3 solicited reactions were reported than expected after the second dose in the adjuvanted vaccine groups. Neutralising and binding antibody responses after two vaccine doses were higher in adjuvanted versus unadjuvanted groups, in AS03-versus AF03-adjuvanted groups, in HD versus LD groups, and in younger versus older age strata.

**Interpretation:** The lower than expected immune responses, especially in the older age stratum, and the higher than anticipated reactogenicity post dose 2 were likely due to a higher than anticipated host cell protein content and lower than planned antigen dose in the clinical material. Further development of the AS03-adjuvanted candidate vaccine will focus on identifying the optimal antigen formulation and dose.

## Introduction

Coronavirus disease of 2019 (Covid-19), caused by the novel coronavirus, severe acute respiratory syndrome coronavirus 2 (SARS-CoV-2), emerged in December 2019 in Wuhan City, China, and spread rapidly to become a Public Health Emergency of International Concern in January 2020 ^1^; a global pandemic was declared in March 2020. The ongoing pandemic has had, and continues to have, catastrophic consequences with over 1.8 million deaths and over 87.3 million confirmed cases to date worldwide (as of 07 January 2021).^2^ Interventions to reduce transmission (including the isolation of affected individuals and those deemed at high-risk of severe outcomes, the use of face masks and limiting face-to-face interactions between individuals from separate households) have been undertaken on an unprecedented worldwide scale and have had far reaching socio-economic impact.^3^ The dominant Covid-19 disease manifestations are fever, fatigue and dry cough.^4,5^ While most young people and children tend to have only mild symptoms or are asymptomatic, adults over age 50 years and individuals with chronic medical conditions are at increased risk of severe outcomes and death.^4,6^

Vaccination against SARS-CoV-2 will likely provide the most effective interventional long-term means of preventing and controlling SARS-CoV-2 infection. The development of safe, effective vaccines against SARS-CoV-2 is therefore an urgent global priority. Of over 60 vaccines currently in clinical development, several have reached Phase III testing,^7^ with interim efficacy results already available for a number of these through peer-reviewed publications^8,9^ or public statements.^10-12^ At the time of writing, the mRNA-based vaccine BNT162b2 (Pfizer, BioNTech) has received conditional marketing authorization, emergency use authorization or a temporary authorization in a number of regions including the USA, EU, UK, Mexico, and Canada (for use in relation to the Covid-19 pandemic); another mRNA vaccine mRNA-1273 (Moderna, Inc.) also received approval for emergency use in the USA and in Europe; and the chimpanzee adenovirus vectored vaccine ChAdOx1 nCoV-19 (AstraZeneca, Oxford University) was approved for emergency use in the UK.

The SARS-CoV-2 spike (S) glycoprotein on the virion surface mediates host cell entry, making the S protein a key target in vaccine development. Previous work on the S protein of the closely-related Middle East respiratory syndrome coronavirus (MERS-CoV), showed that the introduction of double proline substitutions at the beginning of the central helix in the S2 subunit stabilised the protein in a prefusion conformation, preventing the major conformational changes that occur during fusion of the viral and host cell membranes.^13^ This prefusion S protein elicited potent neutralising antibody responses in mice.^13^ Wrapp and colleagues successfully applied this stabilising strategy to the SARS-CoV-2 S protein.^14^ Sanofi Pasteur has developed a candidate SARS-CoV-2 recombinant protein vaccine containing the stabilised SARS-CoV-2 prefusion S protein as the vaccine target, as have other vaccine developers.

Recombinant protein vaccines offer the advantages of fewer potential safety concerns and lower production costs over other traditional vaccines, which rely mostly on attenuation or inactivation of the pathogen.^15^ However, they often require the use of an adjuvant to enhance the magnitude, quality, and persistence of the immune response.^15^ The antigen dose-sparing qualities of a formulation containing an adjuvant allows a reduced quantity of vaccine antigen to achieve a robust immune response compared to antigen alone. This may be of particular importance in a pandemic situation, where there may be potential constraints in antigen supply. We tested two different oil-in-water emulsions as vaccine adjuvant components in the current Phase I/II trial: the AF03 adjuvant (Sanofi Pasteur, France)^16^ and the AS03 Adjuvant System (GlaxoSmithKline, Wavre, Belgium).^17^

This first-in-human study evaluated the safety and immunogenicity, including binding and neutralising antibody responses and cell-mediated immunity, of the candidate vaccine with the goal of informing selection of an adjuvant formulation, antigen dose, and immunisation schedule to proceed to further clinical development.

## Methods

### Study design and participants

This is a Phase I/II, randomised, modified double-blind (observer-blind), first-in-human, parallel group, placebo-controlled, and dose-ranging study (NCT04537208), with a sentinel safety cohort and early safety data review. The study is conducted across 10 centres in the USA, with a planned duration of approximately 12 months following the last study injection. Here, we present interim safety and immunogenicity data up to 43 days (D43) after first vaccination with the stabilised pre-fusion S (preS) protein vaccine, CoV2 preS dTM.

The study was undertaken in compliance with the International Conference on Harmonisation (ICH) guidelines for Good Clinical Practice and the principles of the Declaration of Helsinki. The protocol and amendments were approved by applicable Independent Ethics Committees/Institutional Review Boards and the regulatory agency as per local regulations. Informed consent was obtained from the participants before any study procedures were performed.

Healthy adults aged 18 years and older were eligible for inclusion. A lateral flow immunochromatographic assay (COVID-19 IgG/IgM Rapid Test Cassette, Healgen Scientific, MD, USA) was used to identify those with recent or prior SARS-CoV-2 infection; the test was performed at the clinical site by trained personnel, according to the manufacturer’s instructions. Individuals testing negative for SARS-CoV-2 antibodies were included in the study. Exclusion criteria included chronic illness or medical conditions considered to potentially increase the risk for severe Covid-19 illness; women who were pregnant or lactating; women of childbearing potential who were not using an effective method of contraception or abstinence from at least 4 weeks prior to the first vaccination until at least 12 weeks after the last vaccination; participation, or planned participation, in another clinical trial during the study period; receipt or planned receipt of any vaccine in the 30 days before the first or up to 30 days following the last study vaccination (except for influenza vaccination, which may be received at least 2 weeks before or after study vaccines); receipt of immunoglobulins, blood or blood-derived products in the past 3 months; and active or prior documented autoimmune disease.

Participants were stratified by age (18–49 years and ≥50 years) and randomised to 11 different treatment groups to receive one of five candidate vaccine formulations or placebo, as a single-dose or two-dose schedule. The first injection was on D1 and the second on D22 (**Supplementary Figure S1**). The candidate vaccine formulations were: low-dose (LD) antigen with AF03 or AS03 adjuvant, high-dose (HD) antigen with AF03 or AS03 adjuvant, or unadjuvanted HD antigen (**Supplementary Table S1**). No participants aged 50 years and older were allocated to the unadjuvanted HD antigen group, as older adults are less likely to respond without the presence of an adjuvant and to minimise the theoretical risk of vaccine enhanced disease. Groups were randomly assigned using an interactive response technology system. Block randomisation was used, with blocks of varying sizes, whereby greater numbers of participants were allocated to the two-dose, AS03-adjuvanted groups (**Supplementary Table S1**). Only the study site staff who prepared and administered the vaccine knew which vaccine was administered, and they were not involved in assessment of adverse events or of the study data.

Initially, 30 participants aged 18–49 years were enrolled into a safety sentinel cohort and received a single dose of the intervention to which they were randomised. A review of the safety data up to nine days after the first dose, unblinded to treatment group, was carried out by the Sanofi Pasteur internal safety committee. Only upon demonstration of acceptable safety were the remaining participants enrolled.

A participant subset from the two-dose cohort, across both age strata, was randomly assigned for evaluation of cell-mediated immunity (CMI).

### Vaccines and vaccination

The targeted quantities of the SARS-CoV-2 preS antigen per vaccine dose were 5 µg for the low-dose formulation and 15 µg for the high-dose formulation. However, during characterisation studies on the final bulk drug substance, a key polyclonal antibody reagent used to detect the SARS-CoV-2 preS protein was found to also recognise glycosylated host cell proteins (HCP). As a result, the purity and HCP levels reported for the Phase I/II clinical trial materials were inaccurate and the concentration of SARS-CoV-2 preS protein in the formulated vaccine product was significantly lower (approximately 4–6 fold) than planned. Upon re-calculation, the effective dose levels administered in a 0.5 mL vaccine dose in this study were 1.3 µg (LD) and 2.6 µg (HD) of functional SARS-CoV-2 preS protein. The underdosing of the vaccine formulation was discovered after the study was fully enrolled and all participants had received at least one dose of their assigned product. The differences between the targeted and the effective dose levels correspond to an excess HCP content in the clinical materials (recalculated HCP content, 3.7 µg and 12.4 µg, respectively).

The AF03 (Sanofi Pasteur) and AS03 (GlaxoSmithKline) adjuvants are oil-in-water emulsions. One dose of the AF03 adjuvant emulsion^16^ contained 12.5 mg squalene, 1.85 mg sorbitan monooleate (Dehymuls S SMO™), 2.38 mg Macrogol cetostearyl ether (Kolliphor CS12™) and 2.31 mg mannitol in phosphate-buffered saline (PBS), and was presented in a 0.7 mL monodose vial (single dose, 0.25mL per dose). One dose of the AS03 Adjuvant System^17^ contained 11.86 mg α-tocopherol, 10.69 mg squalene and 4.86 mg polysorbate-80 (Tween^®^80) in PBS, and was presented in a 3.15 mL multidose vial (10 doses, 0.25 mL per dose).

Vaccine formulations were supplied in two separate vials, one vial containing antigen suspension and another containing the adjuvant emulsion or PBS diluent. These were mixed prior to injection to give a final dose volume for injection of 0.5 mL, containing 0.25mL antigen and 0.25mL adjuvant emulsion or PBS diluent. Placebo recipients received 0.5 mL 150 mM NaCl. Vaccine formulations and placebo were prepared by qualified and trained study personnel and administered into the deltoid region of the upper arm by intramuscular injection.

### Safety

The primary objective was to describe the safety profile of the candidate vaccine formulations in all participants. Safety endpoints included immediate unsolicited systemic AEs (occurring within 30 minutes after each dose); solicited injection site reactions (pain, erythema and swelling) and solicited systemic reactions (fever, headache, malaise and myalgia) up to 7 days after each dose; clinical safety laboratory measures; unsolicited AEs up to 21 days after each dose; and medically attended adverse events (MAAEs), serious adverse events (SAEs) and adverse events of special interest (AESIs) are documented throughout the study. AESIs included anaphylactic reactions and potential immune-mediated diseases (pIMDs). pIMDs are a subset of AEs that include autoimmune diseases and other inflammatory and/or neurologic disorders of interest which may or may not have an autoimmune aetiology.^18^ AEs were assessed for intensity (grade 1 to grade 3) and their relationship to the study intervention by the Investigator.

Participants were instructed to contact the site if they experienced symptoms of a Covid-19-like illness, defined by specified clinical symptoms and signs, at any time during the study (see **Supplementary Material** for the pre-defined list of signs and symptoms). Nasopharyngeal swabs were collected as soon as possible after the date of first clinical manifestation to test for the presence of SARS-CoV-2 by nucleic acid amplification test (Abbott RealTime SARS-CoV-2 RT-PCR, USA; available under EUA), in which RNA from the samples was extracted and purified and SARS-CoV-2 specific primers were used.

Clinical safety laboratory parameters were measured eight days after the last dose (on D9 for the single-dose cohort and D30 for the two-dose cohort). Laboratory assessments included serum biochemistry tests, haematology (platelet count, haemoglobin, haematocrit, differential white blood cell count [neutrophils, lymphocytes, monocytes, eosinophils and basophils]) and urine analyses.

### Immunogenicity assessment

The primary immunogenicity objective was to describe the neutralising capacity of vaccine-induced antibodies at D1, D22 and D36 for each study group. SARS-CoV-2 neutralising antibodies were measured in serum samples by microneutralisation assay performed at Sanofi Pasteur Global Clinical Immunology (GCI) Swiftwater, PA, USA, using the SARS-CoV-2 USA-WA1/2020 strain (BEI Resources; catalog# NR-52281). The reduction in SARS-CoV-2 infectivity, compared to that in the control wells, indicated the presence of neutralising antibodies in the serum sample. The 50% neutralisation titre was recorded^19^ (**Supplementary Methods**). Secondary objectives for immunogenicity included a description of the binding antibody profile on D1, D22 and D36 for each study group measured using indirect enzyme-linked immunosorbent assay performed at Nexelis, Laval, Quebec, Canada (ELISA; **Supplementary Methods**), in which the reference standard was human serum with known concentration of anti-S protein IgG antibodies; quantitative results were reported in ELISA Units (EU)/mL.

In an exploratory analysis, neutralising antibody titres were measured in a panel of human convalescent serum (Sanguine Biobank, iSpecimen and PPD). Convalescent samples were obtained from 93 donors between 17 and 47 days following PCR-positive diagnosis of Covid-19. Donors had recovered (with clinical severity ranging from mild to severe), and were asymptomatic at time of sample collection.

### Cell-mediated immunity

Th1/Th2 CMI responses were measured from blood samples obtained at D1, D22, and D36, following *ex vivo* stimulation, using the TruCulture^®^ system (Myriad Biosciences, Austin, TX). Blood samples (1 mL) were drawn directly into the TruCulture tubes, containing 2 mL of buffered media without stimulation (negative control), SARS-CoV-2 S protein for specific stimulation (S 2P-GCN4, GeneArt); or Staphylococcal enterotoxin B plus anti-cluster of differentiation [CD]28 for unspecific stimulation (positive control) (**Supplementary Methods**). Validated cytokine profiling panels were used to evaluate relevant cytokine concentrations: IFN-gamma (IFN-γ), tumor necrosis factor alpha (TNF-α), interleukin (IL)-2, IL-4, IL-5 and IL-13) (**Supplementary Methods**).

### Statistical Analysis

All analyses were descriptive; there was no hypothesis testing. No sample size calculations were performed. Approximately 440 participants were planned to be enrolled in this study (**Supplementary Table S1**), with 300 participants aged 18–49 years (N=20 in each group, except AS03-adjuvanted groups in the two-dose cohort, with 60 participants in each group) and 140 participants aged 50 years or older (N=10 in each group, except in AS03-adjuvanted groups in the two-dose cohort, with 30 participants in each group).

Safety objectives were assessed in the safety analysis set (SafAS), defined as randomised participants who received at least one dose; participants were analysed according to the study treatment received. The full analysis set (FAS) included all randomised participants who received at least one dose; participants were analysed according to the treatment group to which they were randomised. The per-protocol analysis set (PPAS) was defined as the subset of participants from the FAS who met all inclusion/exclusion criteria, who had no protocol deviation and who had negative results in the ELISA and/or neutralisation test at baseline; those who had at least one valid post-dose serology sample within the pre-defined time window were included in the PPAS for immunogenicity (PPAS-IAS). CMI analyses were carried out in the subset of PPAS participants who provided at least one CMI sample within the pre-defined time window (PPAS-CMI).

Primary safety endpoints were summarised by study intervention group. Neutralising antibody profiles were described based on geometric mean antibody titres (GMTs) and 95% confidence intervals (CI). Fold-rises in serum antibody neutralisation titre post-vaccination relative to D1 were calculated, whereby pre-vaccination titres below the lower limit of quantification (LLOQ) were converted to LLOQ/2. The percentage of participants with a 4-fold rise in serum neutralisation titre relative to D1 at D22 and D36 (post-/pre-dose) are presented. Neutralising antibody seroconversion was defined as baseline values below the LLOQ with detectable neutralisation titres above assay LLOQ at D22 and D36.

Binding antibody profiles were described based on S-specific antibody geometric mean concentrations (GMCs) measured at D22 and D36. Antibody concentrations <LLOQ were converted to half LLOQ (**Supplementary Methods**).

The 95% CIs for the GMTs, GMCs and GMT ratios were calculated using normal approximation of log10-transformed titres. The 95% CIs for the proportions were calculated using the Clopper-Pearson method.^20^ The differences in the seroconversion rates between groups were computed along with the 2-sided 95% CIs by the Wilson-Score method without continuity correction.^20^

To characterise T-helper cell polarisation, the pre- (D1) to post-vaccination (D22 or D36) fold cytokine rises were computed by treatment group, and ratios of fold rises for cytokine pairs (IFN-γ/IL-4, IFN-γ/IL-5, and IFN-γ/IL-13; IL-2/IL-4, IL-2/IL-5, and IL-2/IL-13; TNFα/IL-4, TNFα /IL-5, and TNFα /IL-13) were computed (**Supplementary Methods**).

Statistical analyses were performed using SAS^®^ Version 9.4.

### Role of the funding source

Funding was provided by Sanofi Pasteur and the US Government through Biomedical Advanced Research and Development Authority (BARDA) under contract HHSO100201600005I.

## Results

Of 441 participants randomised, two participants aged 50 years or older from the LD + AS03 group (two-dose cohort) were found not to meet eligibility criteria. Thus, 439 participants received at least one dose and were included in the FAS: 269 in the two-dose cohort (N=179 aged 18–49 years and N=90 aged ≥50 years; **Figure 1**) and 170 in the single-dose cohort (N=120 aged 18–49 years and N=50 aged ≥50 years; **Supplementary Figure S2**). Due to an issue in the specifications of the randomisation system, more participants in the younger age stratum were allocated to the single-dose cohort and fewer participants were allocated to the two-dose cohort than planned. The male:female participant ratio was balanced overall, and across most treatment groups. Most enrolled participants were white **(Table 1; Supplementary Table S2)**.

**Table 1.** Participant demographic characteristics (FAS) per treatment group, for participants in the two-dose cohort.

|  | **LD + AF03**<br>**N=28** | **LD + AS03**<br>**N=82** | **HD + AF03**<br>**N=27** | **HD + AS03**<br>**N=85** | **HD (no adjuvant)**<br>**N=18** | **Placebo**<br>**N=29** |
| --- | --- | --- | --- | --- | --- | --- |
| **18-49 years** |  |  |  |  |  |  |
| Female, n (%) | 9/18 (50.0) | 25/54 (46.3) | 11/17 (64.7) | 23/54 (42.6) | 10/18 (55.6) | 8/18 (44.4) |
| Mean age, years (SD) | 35.3 (8.67) | 33.7 (8.99) | 32.5 (9.12) | 34.9 (8.77) | 34.8 (10.2) | 32.0 (8.41) |
| Racial origin, n (%) |  |  |  |  |  |  |
| White | 15/18 (83.3) | 46/54 (85.2) | 16/17 (94.1) | 48/54 (88.9) | 16/18 (88.9) | 16/18 (88.9) |
| Asian | 2/18 (11.1) | 4/54 (7.4) | 0/17 | 4/54 (7.4) | 1/18 (5.6) | 1/18 (5.6) |
| Black or African American | 1/18 (5.6) | 4/54 (7.4) | 1/17 (5.9) | 2/54 (3.7) | 1/18 (5.6) | 1/18 (5.6) |
| American Indian or Alaska Native | 0 | 0 | 0 | 0 | 0 | 0 |
| Ethnicity, n (%) |  |  |  |  |  |  |
| Hispanic or Latino | 4/18 (22.2) | 9/54 (16.7) | 2/17 (11.8) | 6/54 (11.1) | 3/18 (16.7) | 3/18 (16.7) |
| **50 years and older** |  |  |  |  |  |  |
| Female, n/N (%) | 5/10 (50.0) | 11/28 (39.3) | 7/10 (70.0) | 21/31 (67.7) | NA | 6/11 (54.5) |
| Mean age, years (SD) | 58.8 (7.69) | 59.8 (6.53) | 58.7 (7.92) | 61.7 (7.86) | NA | 61.5 (4.41) |
| Racial origin, n/N (%) |  |  |  |  |  |  |
| White | 9/10 (90.0) | 24/28 (85.7) | 9/10 (90.0) | 30/31 (96.8) | NA | 10/11 (90.9) |
| Asian | 1/10 (10.0) | 3/28 (10.7) | 0 | 0 | NA | 0 |
| Black or African American | 0 | 0 | 0 | 1/31 (3.2) | NA | 1/11 (9.1) |
| American Indian or Alaska Native | 0 | 1/28 (3.6) | 0 | 0 | NA | 0 |
| Multiple | 0 | 0 | 1/10 (10.0) | 0 | NA | 0 |
| Ethnicity, n/N (%) |  |  |  |  |  |  |
| Hispanic or Latino | 0 | 2/28 (7.1) | 1/10 (10.0) | 3/31 (9.7) | NA | 0 |

HD, high-dose; LD, low-dose; N, total number of participants; n, number of participants for the specified item

**Figure 1:**
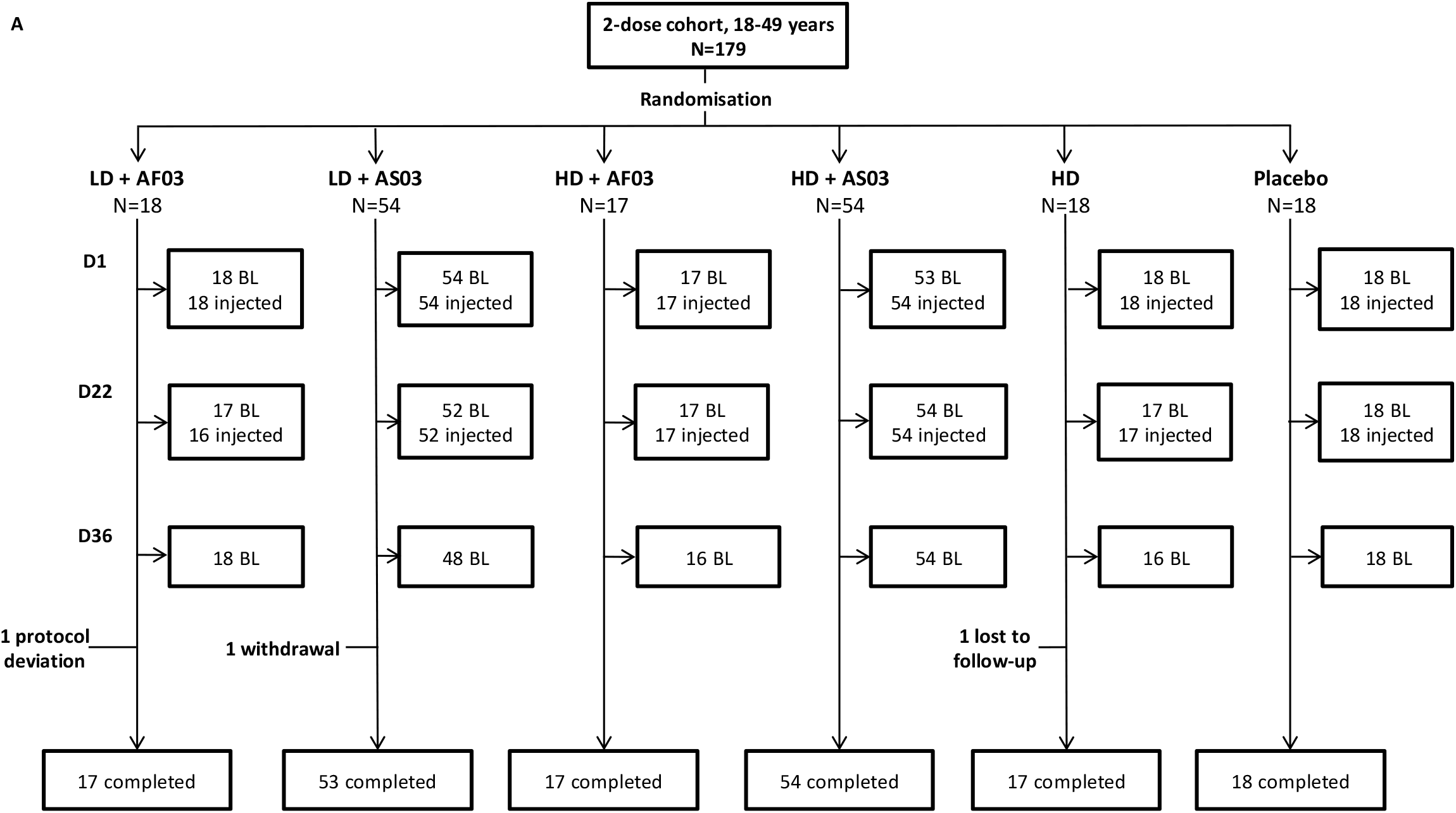
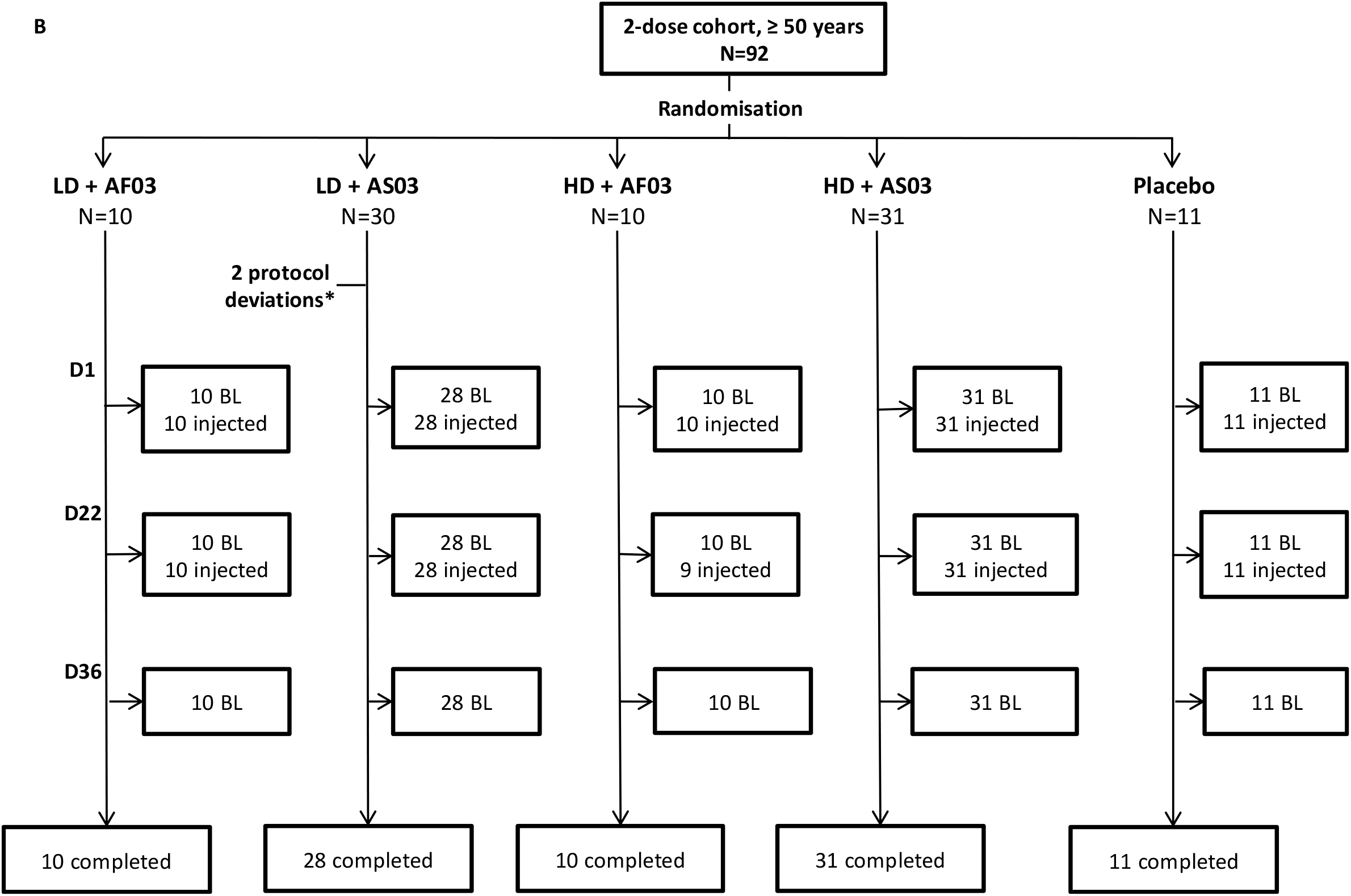
Participant flow through the study for those randomised to receive two vaccine doses in the (A) 18–49 years and (B) ≥ 50 years age strata. *Two participants, who were found not to meet all inclusion criteria after randomisation, were discontinued before receiving the first dose. BL, number of participants for whom blood was collected; LD, low-dose; HD, high-dose; N, number of participants randomised to each treatment group

### Safety

Among the adjuvanted vaccine groups, solicited injection site reactions (pain, erythema and swelling) during the first 7 days after vaccination, including grade 3 reactions, occurred more frequently after the second dose than after the first dose, in both age strata (two-dose cohort; **Figure 2**). The unadjuvanted HD formulation had a low frequency of injection site reactions, similar to the placebo group. Overall, pain was the most frequently reported injection site reaction (post-dose 2, range across adjuvanted vaccine groups, 73.1%–91.8%). Solicited injection site reactions were generally less frequent and less severe in participants aged 50 years and older compared to participants aged 18–49 years across adjuvanted groups, in AF03-adjuvanted formulations compared the AS03-adjuvanted formulations, and in the LD compared to HD formulation (two-dose cohort; **Figure 2**). No grade 3 solicited injection site reactions were observed after the first dose in any treatment group, and none were observed in the unadjuvanted HD or placebo groups after one or two doses. After the second dose, grade 3 injection site reactions occurred more frequently than expected in AS03-adjuvanted formulations, with the highest frequency in the HD + AS03 group; the most frequent grade 3 injection site reaction was erythema (20/85 [23.5%] observed in participants in the HD + AS03 group), followed by swelling (11/85 [12.9%], HD + AS03). No grade 3 injection site reactions were considered serious and all resolved within a median duration of 2 days.

**Figure 2.**
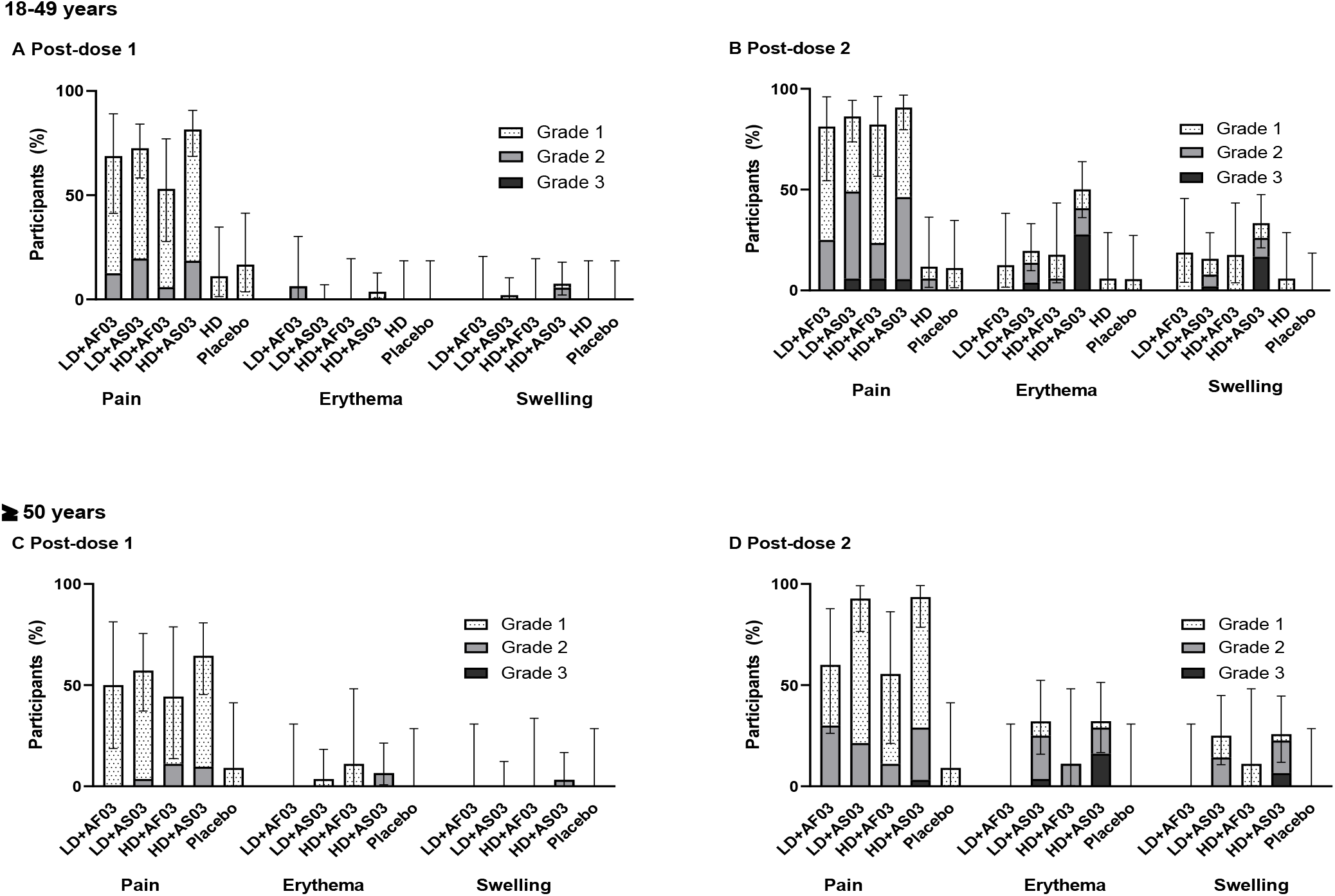
The frequency of solicited injection site reactions after the first or second dose in participants from the two-dose cohort (SafAS).

Among the adjuvanted vaccine groups, solicited systemic reactions (fever, headache, malaise and myalgia) during the first 7 days after vaccination, including grade 3 reactions, were also reported more frequently after the second dose than after the first dose in both age strata (two-dose cohort; **Figure 3**). They were generally less frequent and less severe in participants aged 50 years and older than those in the younger age stratum, and in AF03-adjuvanted formulations compared with the AS03-adjuvanted formulations. The frequency of solicited systemic reactions among the unadjuvanted vaccine and placebo groups was low and similar in both groups (two-dose cohort; **Figure 3**). Myalgia, malaise, and headache were the most commonly reported solicited systemic reactions after the second dose of adjuvanted vaccine (range across adjuvanted vaccine groups: 46.2%–76.5% [myalgia], 50.0%–75.3% [malaise] and 50.0%–71.3% [headache]). Fever was reported after the second dose in 15.4%–35.9% of participants across adjuvanted vaccine groups. Grade 3 systemic reactions also occurred more frequently after the second dose among the adjuvanted formulations, with the highest frequency in the HD + AS03, LD + AS03, and HD + AF03 groups (headache: 9/80 [11.3%], 6/85 [7.1%], and 1/26 [3.8%]; malaise: 13/79 [16.5%], 14/85 [16.5%], and 3/26 [11.5%]; myalgia: 11/80 [13.8%], 9/85 [10.6%], and 3/26 [11.5%], respectively) (**Figure 3**). No Grade 3 solicited systemic reactions were considered serious, and all resolved with a median duration of 2 days.

**Figure 3.**
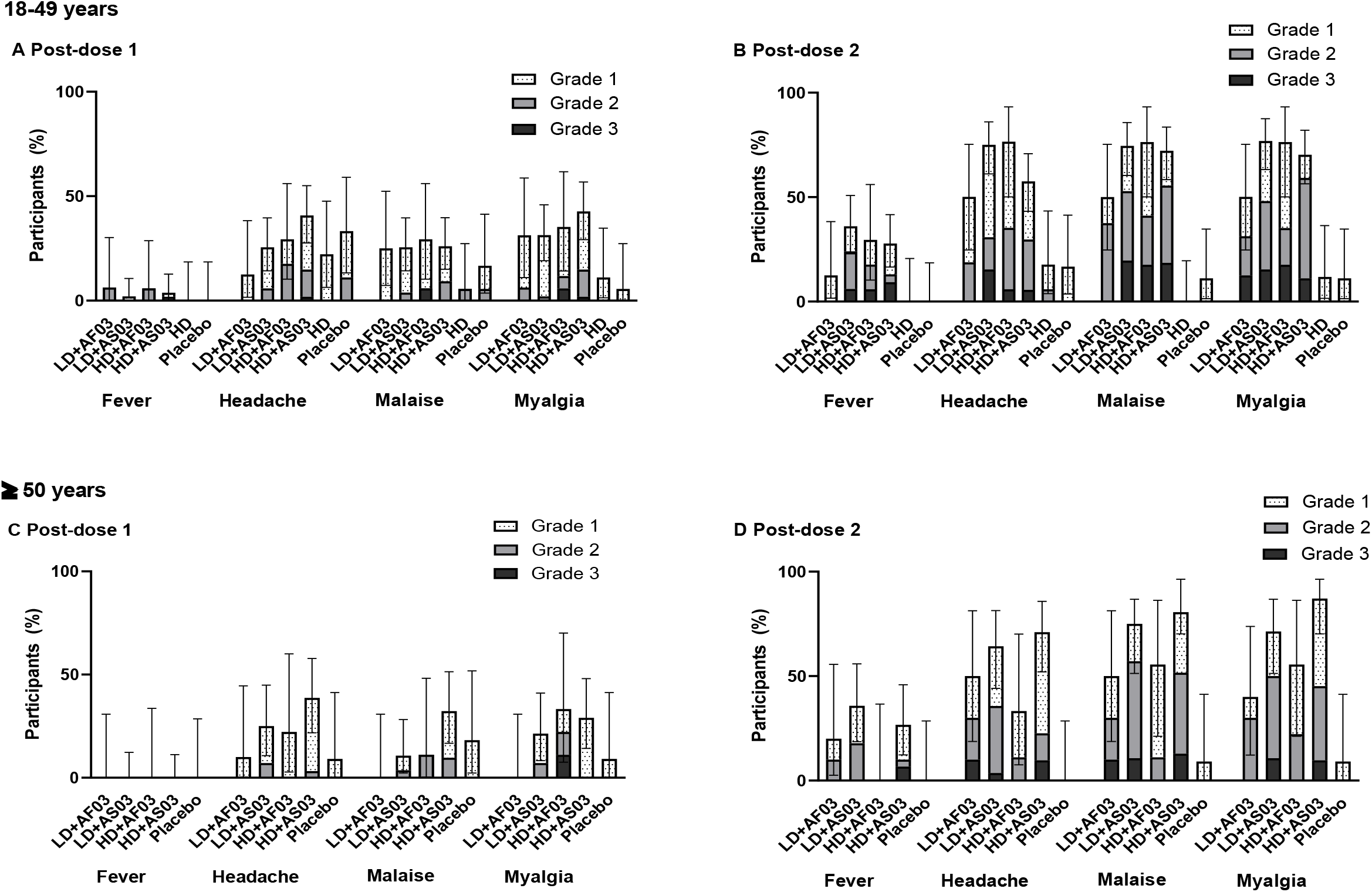
The frequency of solicited systemic reactions after the first or second dose in participants from the two-dose cohort (SafAS)

The reactogenicity profiles (solicited injection site and solicited systemic events) were comparable across the adjuvanted groups in the single-dose cohort (**Supplementary Figure S3**).

No related immediate unsolicited reactions were observed in any group (**Supplementary Table S3**). We observed an increase in the number of unsolicited AEs after the second dose compared to placebo, mainly in the AS03 groups, with a limited number of Grade 3 reactions, primarily due to reporting of reactogenicity-type (local and systemic) events (**Supplementary Table S3**). Three SAEs were reported (two in the single-dose cohort, one each in the LD + AF03 and HD +AF03 groups; and one in the two-dose cohort, in the HD + AS03 group). All 3 were assessed as not related to the vaccine by the Investigator and the Sponsor and none led to discontinuation from the study. These SAEs included one transient ischaemic attack in a participant with a history of ocular occlusion and treatment with antiplatelet agents, who did not receive a second dose; one hip fracture following the first dose in a participant who continued in the study; and one participant who developed breast cancer, who had symptoms prior to study vaccination, and did not receive a second dose. There were no pIMDs reported and there were no AESIs, SAEs or severe MAAEs considered to be related to study vaccine by the Investigator in any group (up to D43 of the study).

### Immunogenicity

A single vaccine dose did not generate neutralising antibody titres above placebo levels in any group at D22 or D36 (**Supplementary Figure S4**). Among participants receiving two doses, neutralising antibody GMTs increased for all adjuvanted vaccine groups by D36, but to a lesser extent among those aged 50 years and older compared to the 18–49 years age stratum (**Figure 4A**). The highest GMTs (95% CI) were observed for the HD + AS03 group (D36: 75.1 [50.5, 112] in the 18–49 years stratum and 52.3 [25.3, 108] for the 50 years and older stratum); the GMT for the pooled convalescent plasma panel was 72.4 (17.6, 297.5) (**Figure 4A**). Post-dose 2, the GMTs observed with the unadjuvanted HD formulation did not differ substantially from those in the placebo group. AF03- and AS03-adjuvanted HD formulations yielded approximately 3-fold and 4-fold higher GMTs than their LD counterparts: 30.2 (16.3, 55.9) and 67.6 (47.9, 95.4) versus 11.7 (6.50, 20.9) and 17.2 (12.1, 24.5) for HD + AF03 and HD + AS03 versus LD + AF03 and LD + AS03, respectively (both age strata combined). At D36, the percentages of participants with a 4-fold rise in neutralising antibody titres were higher in the HD + AS03 group (66.7% [43.0, 85.4]) than in the LD+AS03 group (36.5% [24.7, 49.6]) (**Supplementary Figure S5A**). The percentages of participants with a 4-fold rise in neutralising antibody titres were lower in the older than the younger age stratum, particularly for participants aged 60 years or older (range across vaccine groups for ≥ 60 years, 0% [0, 84.2] in the LD + AF03 group to 50.0% [15.7, 84.3]) in the HD + AS03 group; N=1 and N=8, respectively; (**Supplementary Figure S5A**). Seroconversion for neutralising antibody titres at D36 was also observed in higher percentages of participants receiving the HD + AS03 formulation compared to the LD + AS03 formulation (88.2% [78.1, 94.8] versus 52.4% (39.4, 65.1]) respectively (**Supplementary Figure S5B**), and lower for older participants, particularly those aged 60 years or older, than younger participants (range across vaccine groups for ≥ 60 years, 0% [0, 97.5] in the LD + AF03 group to 62.5% [24.5, 91.5] in the HD + AS03 group; N=1 and N=8, respectively) **Supplementary Figures S5B**).

**Figure 4.**
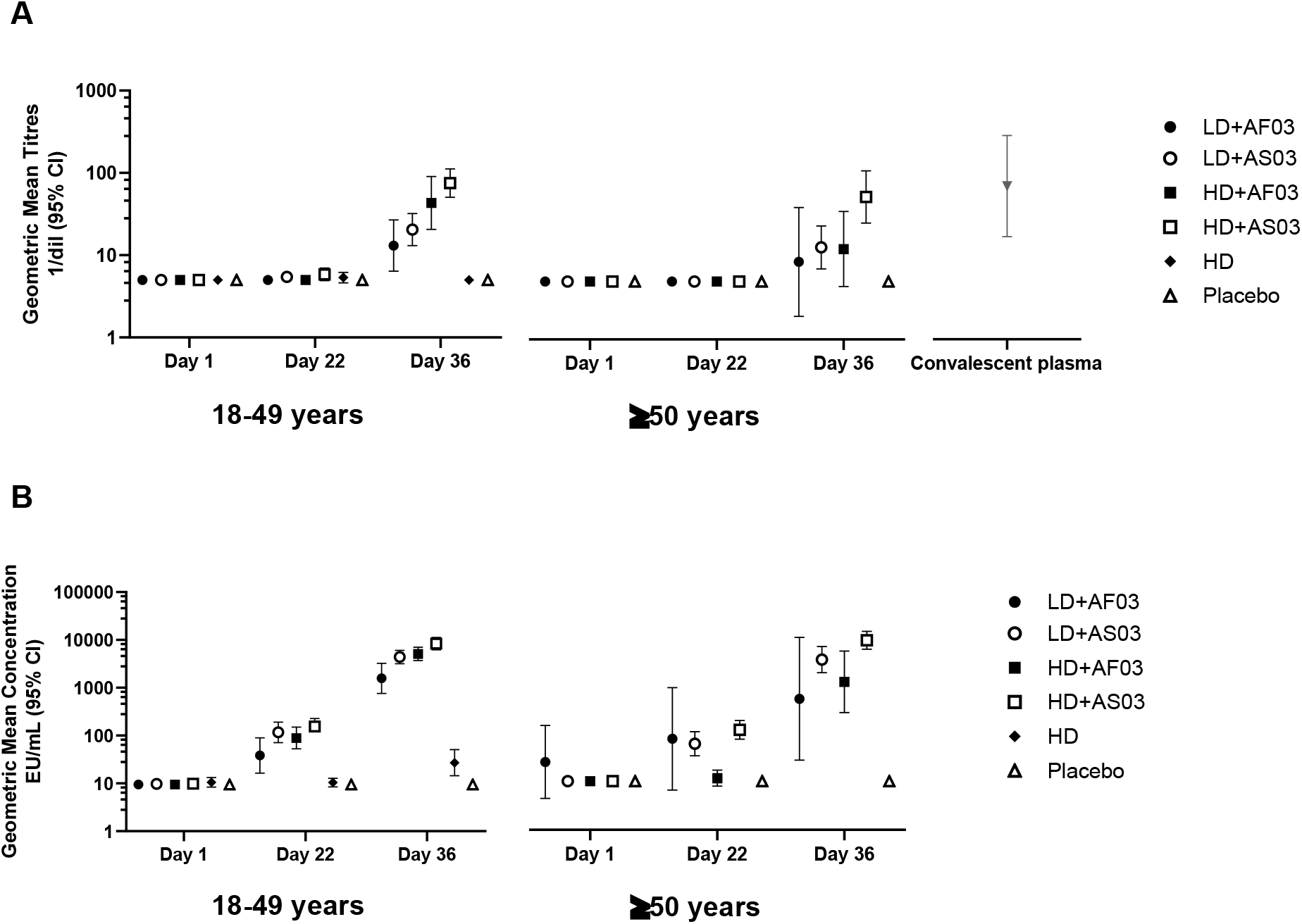
Neutralising antibody titres (microneutralisation assay; A) and binding antibody responses (ELISA; B) after the first and second doses, by age strata (two-dose cohort; PPAS-IAS). Footnote: Neutralising antibody titres are additionally shown in (A) for a panel of 93 convalescent serum samples.

Some increases in binding antibody GMCs could be measured after the 1st dose, but more robust increases were evident after the 2nd dose in both age groups (**Figure 4B**). Responses were higher in the AS03-adjuvanted groups than in the AF03-adjuvanted groups, and higher in the HD groups than the LD groups; a small increase compared with placebo at D36 was observed in the unadjuvanted HD group.

### Cell-mediated immunity

CMI was assessed in a subset of 87 participants from the two-dose cohort: 60 participants aged 18– 49 years (18 per group for the AS03-adjuvanted vaccine; 6 per group for all other study groups) and 27 participants aged ≥50 years and older (9 per group in AS03-adjuvanted groups; 3 per group in all other study groups). Increases of IFN-γ, IL-2 and TNFα cytokines from pre-vaccination to D22 and D36 tended to be more robust compared to the increases for IL-4, IL-5 and IL-13, especially in the AS03-adjuvanted groups, suggesting no Th2 bias in the cell-mediated responses (Supplementary Figure S6).

## Discussion

In this descriptive, first-in-human study, a recombinant, prefusion-stabilised trimeric SARS-CoV-2 S-antigen, formulated with either AS03 or AF03 oil-in-water-based adjuvants, elicited neutralising antibodies with no specific safety concerns that preclude further development. Furthermore, a non-Th2 biased cytokine response was generated in AS03-adjuvanted vaccine groups, with IFN-γ production consistent with previous observations using the AS03 Adjuvant System in influenza, hepatitis B and SARS-CoV-2 candidate vaccines.^21-24^ Two doses of the adjuvanted candidate vaccine formulations were needed to generate a meaningful antibody response in our population of SARS-CoV-2 seronegative individuals. The need for an adjuvant was demonstrated as the unadjuvanted HD group did not elicit a neutralising antibody response. Neutralising antibody titres among participants aged 50 years and older were lower than those in the younger age stratum, demonstrating an age-effect with the formulations evaluated. This was further evidenced by a lower proportion with a 4-fold rise and who seroconverted among older participants, particularly those aged 60 years and older, a key population at risk of severe illness following infection with SARS-CoV-2. The HD formulation given with either AS03 or AF03 resulted in higher neutralising responses than the corresponding LD formulation. The HD + AS03 formulation consistently showed more robust neutralising and binding antibody responses compared to the other candidate vaccine formulations. However, not all participants in the HD + AS03 group achieved seroconversion of neutralising antibodies after the second dose, especially among older participants (62.5% [24.5, 91.5] of those aged ≥60 years).

Previous human experimental coronavirus infection studies identified that the presence of pre-challenge neutralising antibodies was predictive of protection from infection or symptoms following challenge.^25,26^ A growing body of evidence from animal models suggests a key role for humoral responses, and specifically neutralising antibody responses, in protection against SARS-CoV-2. Specifically, a recent investigation showed that adoptive transfer of purified IgG from convalescent macaques protects naïve recipient rhesus macaques against SARS-CoV-2 challenge in a dose-dependent fashion, with relatively low neutralising antibody titres sufficient to protect against SARS-CoV-2 in this model.^27^ High neutralising antibody titres were able to achieve full protection in macaques, whereas 10-fold lower titres could still attain partial protection. These data suggest that neutralising antibodies might be sufficient for protection, even in the absence of cellular and innate immune responses.^27^ Our findings indicate that, while the best performing candidate vaccine formulation among younger adults was comparable in terms of neutralising titres to the titres seen in patients recovering from PCR-confirmed infection (convalescent serum titres), the responses in older participants were notably lower than convalescent serum titres. We therefore conclude that the vaccine candidates tested here have not adequately evaluated the antigen formulation and dose needed to ensure optimal immune responses, including in those most at risk.

The antigen doses evaluated in this trial were substantially lower than the doses initially planned. Further development of the manufacturing process and assays to support characterisation of antigen content were ongoing in parallel to this first-in-human trial. Polyclonal sera used in antigen characterisation were discovered, after the study had commenced, to also bind to HCPs, resulting in an overestimation of the S antigen content during manufacture and underestimation of the HCP content. Although our results support the selection of the AS03 adjuvant for further development, they also indicate that further optimisation of the antigen formulation/purification process and antigen dosage for the selected AS03-adjuvanted candidate vaccine is required.

No related SAEs, related AESIs or related severe MAAEs were reported after either one or two vaccine doses. There was a higher than anticipated number and severity of local and systemic solicited reactions after the second dose of the adjuvanted vaccine formulations, with the highest frequency in the HD + AS03 groups. These reactions were generally less frequent and milder in the older adults than in younger adults. The unadjuvanted HD formulation generated similar reactogenicity profiles to placebo. Overall, these solicited local and systemic reactions were all non-serious, lasted a median of two days and fully resolved. Although transient, the reactogenicity profile observed after the second dose in the adjuvanted groups showed more frequent and more severe reactions compared to those reported in previous studies of influenza vaccine candidates (2-dose schedules) containing the same adjuvants,^28-31^ and compared to the reactogenicity profiles reported for other SARS-CoV-2 vaccine candidates using AS03-adjuvanted recombinant S proteins (Medicago [NCT04450004]^24^ and Clover Biopharmaceuticals [NCT04405908]). We hypothesise that the observation of higher than expected reactogenicity may be explained by the higher than anticipated content of HCP in the clinical material (estimated at approximately 3.7 μg per vaccine dose in the LD groups and 12.4 μg per dose in the HD groups) resulting from the erroneous characterisation of S protein and HCP content. While a high HCP content has been administered historically in the context of clinical development of a recombinant influenza vaccine using the same manufacturing platform, no such levels of HCPs have been previously administered in combination with an adjuvant or in a two-dose injection schedule. In future clinical studies with the CoV2 preS dTM vaccine we therefore plan to utilise clinical material with HCP content below that of the LD group in the present study, with the aim of improving the reactogenicity profile. There was no other medically relevant safety observation during the interim study period (up to D43). Safety monitoring continues for up to 12 months after administration of the second vaccine dose.

A previously postulated theoretical safety concern with SARS-CoV-2 vaccines is the risk of potentiating immunopathology in vaccine recipients upon exposure to wild-type virus.^32^ Various hypothetical risk factors have been proposed, including the magnitude of the immune responses, the balance between binding and functional antibodies, the induction of antibodies with functional characteristics such as binding to particular Fc receptors, and the nature of the T-helper cell response, with Th2-polarised cellular responses being proposed to contribute to immunopathology.^33-36^ In this interim analysis, there was no evidence of vaccine-mediated disease enhancement. The results from our CMI analysis do not support a bias towards Th2 polarisation after the first or second dose of the AS03-adjuvanted candidate vaccine formulations; rather, we observed a consistent Th1 response, as measured through IFN-γ, combined with low levels of Th2 responses, as measured through Il-4, IL-5 and IL-13. While there is currently no evidence of any candidate SARS-CoV-2 vaccine giving rise to the phenomenon of vaccine-mediated enhanced disease, the observed CMI profile is reassuring.

The main limitation of this study was the erroneous characterisation of the content of protein and HCPs used in the filled clinical materials administered in the trial, resulting in a significantly lower concentration of SARS-CoV-2 S protein in the formulated vaccine product than expected, and a correspondingly higher HCP content. Other limitations included the necessarily limited numbers of participants in this Phase I/II study, thus rare SAEs and AESIs may not have been captured. Cellular profiling would also be required to define the source of cytokines detected during *ex vivo* whole blood stimulation.

The results from the candidate vaccine formulations tested here are informative in terms of the neutralising and binding antibody responses generated in healthy adults. Further improvement of the preS vaccine antigen formulation is needed in order to be able to identify the optimal vaccine dose for larger scale Phase III development.

## Supporting information

VAT01_Supplementary Materials_MedRxiv

## Data Availability

Qualified researchers may request access to patient level data and related study documents including the clinical study report, study protocol with any amendments, blank case report form, statistical analysis plan, and dataset specifications. Patient level data will be anonymised and study documents will be redacted to protect the privacy of trial participants. Further details on Sanofi data sharing criteria, eligible studies, and process for requesting access can be found at: https://www.clinicalstudydatarequest.com.

## Contributors

BF, ALC, MIB, CAD and GDB contributed to the concept or design of the study; PAG, MGD, BJE, IF, HJ, MCK, MAK, RM, HS, LDS and JS contributed to data acquisition; and PAG, BF, ALC, MIB, OH, SJS, LS, FTDS, MK, SG, CAD and GDB were involved in the analysis and/or interpretation of the data. All authors were involved in drafting or critically revising the manuscript, and all authors approved the final version and are accountable for the accuracy and integrity of the manuscript.

## Declaration of interests

BF, ALC, MIB, OH, HJ, RM, SJS, JS, SG, CAD and GDB are Sanofi Pasteur employees and may hold stock. MK, LS and FTDS are employed by the GlaxoSmithKline (GSK) group of companies. MK, LS and FTDS hold restricted shares in the GSK group of companies. IF reports grants from Janssen and personal fees from Gilead and ViiV/ GlaxoSmithKline. PAG, BJE, MCK, MAK, MGD, LDS and HS declare that they have no conflict of interest.

## Data sharing

Qualified researchers may request access to patient level data and related study documents including the clinical study report, study protocol with any amendments, blank case report form, statistical analysis plan, and dataset specifications. Patient level data will be anonymised and study documents will be redacted to protect the privacy of trial participants. Further details on Sanofi’s data sharing criteria, eligible studies, and process for requesting access can be found at: https://www.clinicalstudydatarequest.com.

## Acknowledgements

The authors would like to thank all participants, investigators and study site personnel who took part in this study. The authors acknowledge Juliette Gray of inScience Communications, Springer Healthcare Ltd, London, UK for editorial assistance with the preparation of this manuscript, funded by Sanofi Pasteur. The authors also thank Isabel Grégoire for editorial assistance and manuscript coordination on behalf of Sanofi Pasteur.

## Notes

**Funding** Sanofi Pasteur and Biomedical Advanced Research and Development Authority (BARDA)

### Clinical Trial

NCT04537208

### Funding Statement

Funding was provided Sanofi Pasteur and by the US Government through Biomedical Advanced Research and Development Authority (BARDA) under contract HHSO100201600005I.

### Author Declarations

https://www.advarra.com/ Pro00045646 approval letter dated 31Jul2020.

## References

1. World Health Organization. Statement on the second meeting of the International Health Regulations (2005) Emergency Committee regarding the outbreak of novel coronavirus (2019-nCoV). https://www.who.int/news/item/30-01-2020-statement-on-the-second-meeting-of-the-international-health-regulations-(2005)-emergency-committee-regarding-the-outbreak-of-novel-coronavirus-(2019-ncov). Accessed 26 October 2020. 2020.

2. Dong E, Du H, Gardner L. An interactive web-based dashboard to track COVID-19 in real time. The Lancet Infectious Diseases 2020; 20(5): 533–4.

3. Nicola M, Alsafi Z, Sohrabi C, et al. The socio-economic implications of the coronavirus pandemic (COVID-19): A review. Int J Surg 2020; 78: 185–93.

4. Guan WJ, Ni ZY, Hu Y, et al. Clinical Characteristics of Coronavirus Disease 2019 in China. The New England journal of medicine 2020; 382(18): 1708–20.

5. Richardson S, Hirsch JS, Narasimhan M, et al. Presenting Characteristics, Comorbidities, and Outcomes Among 5700 Patients Hospitalized With COVID-19 in the New York City Area. Jama 2020; 323(20): 2052–9.

6. Hu B, Guo H, Zhou P, Shi Z-L. Characteristics of SARS-CoV-2 and COVID-19. Nat Rev Microbiol 2020: 1–14.

7. World Health Organization. Draft landscape of COVID-19 candidate vaccines. https://www.who.int/publications/m/item/draft-landscape-of-covid-19-candidate-vaccines. Accessed 26 October 2020. 2020.

8. Polack FP, Thomas SJ, Kitchin N, et al. Safety and Efficacy of the BNT162b2 mRNA Covid-19 Vaccine. The New England journal of medicine 2020.

9. Voysey M, Clemens SAC, Madhi SA, et al. Safety and efficacy of the ChAdOx1 nCoV-19 vaccine (AZD1222) against SARS-CoV-2: an interim analysis of four randomised controlled trials in Brazil, South Africa, and the UK. Lancet 2020.

10. FDA Briefing Document. Moderna COVID-19 Vaccine. Vaccines and Related Biological Products Advisory Committee Meeting December 17, 2020. Available at https://www.fda.gov/media/144434/download. Accessed 28 December 2020.

11. Sinopharm - Beijng Institute of Biological Products. Press Release [Chinese]. Available at http://www.bjbpi.com/news_list.asp?id=787. Accessed 07 January 2021.

12. The Gamaleya National Centre. The first interim data analysis of the Sputnik V vaccine against COVID-19 phase III clinical trials in the Russian Federation demonstrated 92% efficacy. https://sputnikvaccine.com/newsroom/pressreleases/the-first-interim-data-analysis-of-the-sputnik-v-vaccine-against-covid-19-phase-iii-clinical-trials-/. Accessed 07 January 2021.

13. Pallesen J, Wang N, Corbett KS, et al. Immunogenicity and structures of a rationally designed prefusion MERS-CoV spike antigen. Proceedings of the National Academy of Sciences of the United States of America 2017; 114(35): E7348–e57.

14. Wrapp D, Wang N, Corbett KS, et al. Cryo-EM structure of the 2019-nCoV spike in the prefusion conformation. Science (New York, NY) 2020; 367(6483): 1260–3.

15. Nascimento IP, Leite LCC. Recombinant vaccines and the development of new vaccine strategies. Braz J Med Biol Res 2012; 45(12): 1102–11.

16. Klucker MF, Dalençon F, Probeck P, Haensler J. AF03, an alternative squalene emulsion-based vaccine adjuvant prepared by a phase inversion temperature method. Journal of pharmaceutical sciences 2012; 101(12): 4490–500.

17. Garçon N, Vaughn DW, Didierlaurent AM. Development and evaluation of AS03, an Adjuvant System containing α-tocopherol and squalene in an oil-in-water emulsion. Expert review of vaccines 2012; 11(3): 349–66.

18. Tavares Da Silva F, De Keyser F, Lambert PH, Robinson WH, Westhovens R, Sindic C. Optimal approaches to data collection and analysis of potential immune mediated disorders in clinical trials of new vaccines. Vaccine 2013; 31(14): 1870–6.

19. Kalnin KV, Plitnik T, Kishko M, et al. Immunogenicity of novel mRNA COVID-19 vaccine MRT5500 in mice and 2 non-human primates [pre-print]. bioRxiv preprint server 2020: https://doi.org/10.1101/2020.10.14.337535.

20. Newcombe RG. Two-sided confidence intervals for the single proportion: comparison of seven methods. Stat Med 1998; 17(8): 857–72.

21. Diez-Domingo J, Baldo JM, Planelles-Catarino MV, et al. Phase II, randomized, open, controlled study of AS03-adjuvanted H5N1 pre-pandemic influenza vaccine in children aged 3 to 9 years: follow-up of safety and immunogenicity persistence at 24 months post-vaccination. Influenza Other Respir Viruses 2015; 9(2): 68–77.

22. Leroux-Roels G, Marchant A, Levy J, et al. Impact of adjuvants on CD4(+) T cell and B cell responses to a protein antigen vaccine: Results from a phase II, randomized, multicenter trial. Clin Immunol 2016; 169: 16–27.

23. Moris P, van der Most R, Leroux-Roels I, et al. H5N1 influenza vaccine formulated with AS03 A induces strong cross-reactive and polyfunctional CD4 T-cell responses. J Clin Immunol 2011; 31(3): 443–54.

24. Ward BJ, Gobeil P, Séguin A, et al. Phase 1 trial of a Candidate Recombinant Virus-Like Particle Vaccine for Covid-19 Disease Produced in Plants. medRxiv 2020: 2020.11.04.20226282.

25. Callow KA. Effect of specific humoral immunity and some non-specific factors on resistance of volunteers to respiratory coronavirus infection. J Hyg (Lond) 1985; 95(1): 173–89.

26. Reed SE. The behaviour of recent isolates of human respiratory coronavirus in vitro and in volunteers: evidence of heterogeneity among 229E-related strains. J Med Virol 1984; 13(2): 179–92.

27. McMahan K, Yu J, Mercado NB, et al. Correlates of protection against SARS-CoV-2 in rhesus macaques. Nature 2020.

28. Cohet C, van der Most R, Bauchau V, et al. Safety of AS03-adjuvanted influenza vaccines: A review of the evidence. Vaccine 2019; 37(23): 3006–21.

29. Jackson LA, Campbell JD, Frey SE, et al. Effect of Varying Doses of a Monovalent H7N9 Influenza Vaccine With and Without AS03 and MF59 Adjuvants on Immune Response: A Randomized Clinical Trial. Jama 2015; 314(3): 237–46.

30. Levie K, Leroux-Roels I, Hoppenbrouwers K, et al. An adjuvanted, low-dose, pandemic influenza A (H5N1) vaccine candidate is safe, immunogenic, and induces cross-reactive immune responses in healthy adults. The Journal of infectious diseases 2008; 198(5): 642–9.

31. Vesikari T, Pepin S, Kusters I, Hoffenbach A, Denis M. Assessment of squalene adjuvanted and non-adjuvanted vaccines against pandemic H1N1 influenza in children 6 months to 17 years of age. Hum Vaccin Immunother 2012; 8(9): 1283–92.

32. Smatti MK, Al Thani AA, Yassine HM. Viral-Induced Enhanced Disease Illness. Front Microbiol 2018; 9: 2991.

33. Czub M, Weingartl H, Czub S, He R, Cao J. Evaluation of modified vaccinia virus Ankara based recombinant SARS vaccine in ferrets. Vaccine 2005; 23(17-18): 2273–9.

34. Lambert P-H, Ambrosino DM, Andersen SR, et al. Consensus summary report for CEPI/BC March 12-13, 2020 meeting: Assessment of risk of disease enhancement with COVID-19 vaccines. Vaccine 2020; 38(31): 4783–91.

35. Tseng CT, Sbrana E, Iwata-Yoshikawa N, et al. Immunization with SARS coronavirus vaccines leads to pulmonary immunopathology on challenge with the SARS virus. PLoS One 2012; 7(4): e35421.

36. Yasui F, Kai C, Kitabatake M, et al. Prior immunization with severe acute respiratory syndrome (SARS)-associated coronavirus (SARS-CoV) nucleocapsid protein causes severe pneumonia in mice infected with SARS-CoV. Journal of immunology (Baltimore, Md : 1950) 2008; 181(9): 6337–48.

