## Supplementary material for "Safety and immunogenicity of SARS-CoV-2 recombinant protein vaccine formulations in healthy adults: a randomised, placebo-controlled, dose-ranging study": VAT01_Supplementary Materials_MedRxiv

### Supplementary Methods

#### COVID-19-like illness symptoms

Symptoms of COVID-19-like illness are as listed below.

Any ONE of the following (that persist for a period of at least 12 hours or reoccur within a 12-hour period):

- Cough (dry or productive)
- Fever (≥38°C)
- Anosmia (loss of smell)
- Ageusia (loss of taste)
- Chillblains (COVID-toes)
- Difficulty breathing or shortness of breath
- Clinical or radiographic evidence of pneumonia
- Any hospitalization with the following clinical diagnosis:
  - Stroke, myocarditis, myocardial infarction
  - Thromboembolic event (blood clots [eg, pulmonary embolism, deep vein thrombosis, stroke])
  - Purpura fulminans (purplish skin rash)

OR

Any TWO of the following (that persist for a period of at least 12 hours or reoccur within a 12-hour period):

- Pharyngitis
- Chills
- Myalgia
- Headache
- Rhinorrhoea
- Abdominal pain
- At least one of nausea, diarrhoea, vomiting

### Supplementary Methods - Immunogenicity Analyses

#### Microneutralisation assay

The microneutralisation assay was performed at Sanofi Pasteur Global Clinical Immunology, Swiftwater, PA, USA. Serial two-fold dilutions of heat inactivated serum samples (56°C for 30 minutes) were incubated with a challenge dose targeting 50% tissue culture infectious dose ($TCID_{50}$) of SARS-CoV-2 (strain USA-WA1/2020 [BEI Resources; catalog# NR-52281]) at 37°C with 5% $CO_2$ for 1 hour. The serum-virus mixtures were inoculated into wells of a 96-well microplate with preformed Vero E6 (ATCC® CRL-1586™) cell monolayers and adsorbed at 37°C with 5% $CO_2$ for 0.5 hour. Additional assay media was added to all wells without removing the existing inoculum and incubated at 37°C with 5% $CO_2$ for 2 days. After washing and fixation of the Vero E6 cell monolayers, SARS-CoV-2 antigen production in cells was detected by successive incubations with an anti-SARS-CoV nucleoprotein mouse monoclonal antibody (Sino Biological catalog# 40143-MM05), HRP IgG conjugate (Jackson ImmunoResearch Laboratories, catalog #115-035-062), and SeraCare SureBlue® TMB substrate. The resulting optical density (OD) was measured at 450/630 nm using a Molecular Devices microplate reader. The reduction in SARS-CoV-2 infectivity, as compared to that in the virus control wells, constitutes a positive neutralisation reaction indicating the presence of neutralising antibodies in the serum sample. The 50% neutralisation titre ($MN_{50}$) was defined as the reciprocal of the serum dilution for which the virus infectivity was reduced by 50% relative to the virus control on each plate. The $MN_{50}$ for each sample was interpolated by calculating the slope and intercept using the last dilution with an OD below the 50% neutralisation point and the first dilution with an OD above the 50% neutralisation point; $MN_{50}$ titre = (OD of 50% neutralisation point - intercept)/slope. The Lower Limit of Quantitation (LLOQ) of the assay was a titer of 10.

#### Enzyme-linked immunosorbent assay

The SARS-CoV-2 anti-S protein IgG antibodies were measured using an indirect ELISA performed at Nexelis, Laval, Quebec, Canada. Microtitre plates were coated with recombinant SARS-CoV-2 S protein antigen (SARS-CoV-2 S-antigen with 2P mutations in pre-fusion trimer conformation produced in HEK293 cell line, GeneArt, Regensburg, Germany) diluted in coating buffer and incubated overnight (17–21 hours) at 2–8°C. The plates were blocked following the addition of 5% Skim Milk in 1X PBS+ 0.05% Tween-20 blocking buffer to all wells, then incubated for 1 hour at room temperature. Following incubation, the plates were washed with 1X PBS + 0.05% Tween-20 (PBST). The pre-diluted controls, reference, and test samples were then serially diluted in the wells of the pre-coated test plates. Following incubation for 1 hour at room temperature, the plates were washed with PBST, and goat anti-human IgG HRP enzyme conjugate was added to all wells. The plates were incubated for 1 hour at room temperature, washed with PBST and TMB substrate solution was added to all wells. The plates were incubated for substrate development for 30 minutes, stopped by the addition of a stop solution (2N $H_2SO_4$) to all wells. The OD of the microtitre plates were read at 450/620 nm using an assay specific SoftMax Pro template. The average OD value for the plate blank was then subtracted from the ODs within each plate. The sample IgG concentrations were then extrapolated for the blanks, controls, and test samples based on the reference standard curve, which was included on each assay plate within a run. The LLOQ of the assay was established at 18.9 EU/ml.

#### CMI analyses

The TruCulture OptiMAP™ assay (Myriad RBM) is microsphere-based and consists of using antigen-specific antibodies optimized in a capture-sandwich format. All incubations were performed at room temperature. Five μL of a diluted mixture of capture-antibody microspheres were mixed with 5 μL blocker and 10 μL standard, pre-diluted sample, or control in a hard-bottom microtitre plate. TruCulture samples were diluted to the appropriate dilution. The plate was incubated for 1 hour. Ten μL biotinylated detection antibody was added to each well, thoroughly mixed, and incubated for 1 hour. Ten μL diluted Streptavidin-phycoerythrin was added to each well, thoroughly mixed, and incubated for 60 minutes. A filter-membrane microtitre plate was pre-wetted by adding 100 μL wash buffer followed by aspiration via a vacuum manifold device. The reaction contents of the hard-bottom plate were then transferred to the respective wells of the filter plate. All wells were vacuum aspirated, and the contents were washed twice with 100 μL wash buffer. After the last wash, 100 μL wash buffer was added to each well, and the washed microspheres were resuspended with thorough mixing. The plate was then analysed on a Luminex platform. Cytokine concentrations were automatically calculated by adapted software (Myriad RBM) using a standard curve. GM-CSF, IFN-γ, IL-2, IL-4, IL-5, IL-6, IL-10, IL-13, IL-17, TNFα were acquired initially and data for IFN-γ, IL-2, and TNFα are analysed here. IFN-γ is considered the most relevant Th1 polarising cytokine. IL-2 and TNFα are also commonly classified as Th1 cytokines, although they have pleiotropic effects.

*Computation of cytokine fold-rise ratios*

Specific cytokine production was firstly calculated at each timepoint by subtracting cytokine concentration measured in negative control (N) samples from antigen stimulated samples (S). The fold-rise for the individual cytokines at D36/D1 and D22/D1 was calculated by dividing the (S-N) measurement post-vaccination (D22 or D36) by the pre-vaccination (D1) measurement; the fold-rise for individual cytokines were then used to compute the fold-rise ratios and 95% CI for cytokine pairs.

**Supplementary Table S1. Planned group sizes allocated to each treatment group**

| **Single-dose cohort** | | | | |
|---|---|---|---|---|
| | **Dose Level*** | **Adjuvant** | **Number of participants** | |
| | | | **Sentinel Safety† (N=30)** | **Total (N=150)** |
| **18–49 years** | LD | AF03 | 6 | 20 |
| | LD | AS03 | 6 | 20 |
| | HD | AF03 | 6 | 20 |
| | HD | AS03 | 6 | 20 |
| | Placebo | None | 6 | 20 |
| **50 years of age and older** | LD | AF03 | N/A | 10 |
| | LD | AS03 | N/A | 10 |
| | HD | AF03 | N/A | 10 |
| | HD | AS03 | N/A | 10 |
| | Placebo | None | N/A | 10 |

| **Two-dose cohort** | | | |
|---|---|---|---|
| | **Dose Level*** | **Adjuvant** | **Number of participants (N=290)** |
| **18–49 years** | LD | AF03 | 20 |
| | LD | AS03 | 60 |
| | HD | AF03 | 20 |
| | HD | AS03 | 60 |

| | | | |
|---|---|---|---|
| | HD | None | 20 |
| | Placebo | None | 20 |
| **50 years of age and older** | LD | AF03 | 10 |
| | LD | AS03 | 30 |
| | HD | AF03 | 10 |
| | HD | AS03 | 30 |
| | Placebo | None | 10 |

HD, high dose; LD, low dose; N, total participants; N/A, not available (participants aged ≥ 50 years were not allocated to this study arm).

*The targeted dose levels were not met; effective dose levels were 1.3 µg (low-dose) and 2.6 µg (high-dose) of functional vaccine antigen, CoV2 preS, instead of 5 and 15ug, respectively.

†Sentinel safety cohort: Participants in the 18–49 years age stratum from the single-dose cohort were included in the sentinel safety cohort.

Note: A subset of 87 participants in the two-dose cohort (60 participants 18–49 years of age [18 per group in AS03-adjuvanted vaccines; 6 per group in all other study groups]) and 27 participants ≥ 50 years of age [9 per group in AS03-adjuvanted groups; 3 per group in all other study groups, with the exception of the unadjuvanted group]), were randomly assigned to a cellular-mediated immune (CMI) subset.

**Supplementary Table S2. Demographic characteristics of participants in in the single-dose cohort (FAS)**

| | LD + AF03 | LD + AS03 | HD + AF03 | HD + AS03 | Placebo |
|---|---|---|---|---|---|
| 18–49 years | | | | | |
| Female, n/N (%) | 14/24 (58.3) | 9/24 (37.5) | 13/24 (54.2) | 14/24 (58.3) | 12/24 (50.0) |
| Mean age, years (SD) | 36.0 (10.0) | 35.4 (8.66) | 32.0 (9.69) | 29.7 (8.88) | 31.1 (8.45) |
| Racial origin, n/N (%) | | | | | |
| White | 20/24 (83.3) | 21/24 (87.5) | 22/24 (91.7) | 16/24 (66.7) | 20/24 (83.3) |
| Asian | 1/24 (4.2) | 2/24 (8.3) | 1/24 (4.2) | 4/24 (16.7) | 1/24 (4.2) |
| American Indian or Alaska Native | 0/24 | 0/24 | 0/24 | 1/24 (4.2) | 1/24 (4.2) |
| Black or African American | 1/24 (4.2) | 0/24 | 0/24 | 1/24 (4.2) | 0/24 |
| Multiple | 0/24 | 1/24 (4.2) | 1/24 (4.2) | 0/24 | 1/24 (4.2) |
| Not Reported or unknown | 2/24 (8.3) | 0/24 | 0/24 | 2/24 (8.3) | 1/24 (4.2) |
| Ethnicity, n/N (%) | | | | | |
| Hispanic or Latino | 6/24 (25.0) | 2/24 (8.3)* | 3/24 (12.5) | 3/24 (12.5) | 7/24 (29.2) |
| 50 years and older | | | | | |
| Female, n/N (%) | 8/10 (80.0) | 6/10 (60.0) | 4/10 (40.0) | 7/10 (70.0) | 6/10 (60.0) |
| Mean age, years (SD) | 59.3 (7.07) | 57.3 (5.70) | 58.5 (9.29) | 61.3 (10.1) | 60.1 (7.74) |
| Racial origin, n/N (%) | | | | | |
| White | 9/10 (90.0) | 7/10 (70.0) | 8/10 (80.0) | 9/10 (90.0) | 10/10 (100) |
| Asian | 1/10 (10.0) | 1/10 (10.0) | 0/10 | 1/10 (10.0) | 0/10 |
| Black or African American | 0/10 | 1/10 (10.0) | 0/10 | 0/10 | 0/10 |
| American Indian or Alaska Native | 0/10 | 0/10 | 0/10 | 0/10 | 0/10 |
| Native Hawaiian or Other Pacific Islander | 0/10 | 1/10 (10.0) | 0/10 | 0/10 | 0/10 |
| Multiple | 0/10 | 0/10 | 1/10 (10.0) | 0/10 | 0/10 |
| Not Reported or unknown | 0/10 | 0/10 | 1/10 (10.0) | 0/10 | 0/10 |

*Hispanic or Latino ethnicity was not reported for one participant in the HD + AF03 group, in the 18-49 years stratum.
HD, high dose; LD, low dose; N, total participants for that treatment group; n, number of participants for the specific characteristic; SD, standard deviation

### Supplementary Table S3. Summary of unsolicited adverse events within 21 days after any dose for both age strata combined within (A) the two-dose cohort and (B) the single-dose cohort (SafAS).

#### A. Two-dose cohort

| | LD + AF03<br>N=26 | | | LD + AS03<br>N=80 | | | HD + AF03<br>N=26 | | | HD + AS03<br>N=85 | | | HD<br>N=18 | | | Placebo<br>N=29 | | |
|---|---|---|---|---|---|---|---|---|---|---|---|---|---|---|---|---|---|---|
| | **n** | **% (95% CI)** | **AEs** | **n** | **% (95% CI)** | **AEs** | **n** | **% (95% CI)** | **AEs** | **n** | **% (95% CI)** | **AEs** | **n** | **% (95% CI)** | **AEs** | **n** | **% (95% CI)** | **AEs** |
| Unsolicited AE | 12 | 46.2 (26.6; 66.6) | 29 | 34 | 42.5 (31.5; 54.1) | 79 | 11 | 42.3 (23.4; 63.1) | 33 | 50 | 58.8 (47.6; 69.4) | 108 | 5 | 27.8 (9.7; 53.5) | 9 | 9 | 31.0 (15.3; 50.8) | 13 |
| Grade 3 | 1 | 3.8 (0.1; 19.6) | 5 | 4 | 5.0 (1.4; 12.3) | 5 | 1 | 3.8 (0.1; 19.6) | 2 | 3 | 3.5 (0.7; 10.0) | 6 | 1 | 5.6 (0.1; 27.3) | 1 | 0 | 0 (0; 11.9) | 0 |
| Unsolicited AR | 6 | 23.1 (9.0; 43.6) | 6 | 21 | 26.3 (17.0; 37.3) | 42 | 5 | 19.2 (6.6; 39.4) | 9 | 37 | 43.5 (32.8; 54.7) | 70 | 1 | 5.6 (0.1; 27.3) | 1 | 3 | 10.3 (2.2; 27.4) | 4 |
| Grade 3 | 0 | 0 (0; 13.2) | 0 | 1 | 1.3 (0; 6.8) | 1 | 0 | 0 (0; 13.2) | 0 | 3 | 3.5 (0.7; 10.0) | 6 | 0 | 0 (0; 18.5) | 0 | 0 | 0 (0; 11.9) | 0 |
| Unsolicited injection site AR | 3 | 11.5 (2.4; 30.2) | 3 | 6 | 7.5 (2.8; 15.6) | 7 | 2 | 7.7 (0.9; 25.1) | 2 | 20 | 23.5 (15.0; 34.0) | 24 | 1 | 5.6 (0.1; 27.3) | 1 | 1 | 3.4 (0.1; 17.8) | 1 |
| Grade 3 | 0 | 0 (0; 13.2) | 0 | 0 | 0 (0; 4.5) | 0 | 0 | 0 (0; 13.2) | 0 | 0 | 0 (0; 4.2) | 0 | 0 | 0 (0; 18.5) | 0 | 0 | 0 (0; 11.9) | 0 |
| Unsolicited systemic AE | 11 | 42.3 (23.4; 63.1) | 26 | 32 | 40.0 (29.2; 51.6) | 72 | 11 | 42.3 (23.4; 63.1) | 31 | 42 | 49.4 (38.4; 60.5) | 84 | 5 | 27.8 (9.7; 53.5) | 8 | 8 | 27.6 (12.7; 47.2) | 12 |
| Grade 3 | 1 | 3.8 (0.1; 19.6) | 5 | 4 | 5.0 (1.4; 12.3) | 5 | 1 | 3.8 (0.1; 19.6) | 2 | 3 | 3.5 (0.7; 10.0) | 6 | 1 | 5.6 (0.1; 27.3) | 1 | 0 | 0 (0; 11.9) | 0 |
| Unsolicited systemic AR | 3 | 11.5 (2.4; 30.2) | 3 | 18 | 22.5 (13.9; 33.2) | 35 | 4 | (4.4; 34.9) | 7 | 26 | 30.6 (21.0; 41.5) | 46 | 0 | 0 (0; 18.5) | 0 | 2 | 6.9 (0.8; 22.8) | 3 |
| Grade 3 | 0 | 0 (0; 13.2) | 0 | 1 | 1.3 (0; 6.8) | 1 | 0 | (0; 13.2) | 0 | 3 | 3.5 (0.7; 10.0) | 6 | 0 | 0 (0; 18.5) | 0 | 0 | 0 (0; 11.9) | 0 |
| SAE | 0 | 0 (0; 13.2) | 0 | 0 | 0 (0; 4.5) | 0 | 0 | (0; 13.2) | 0 | 1 | 1.2 (0; 6.4) | 1 | 0 | 0 (0; 18.5) | 0 | 0 | 0 (0; 11.9) | 0 |
| Grade 3 | 0 | 0 (0; 13.2) | 0 | 0 | 0 (0; 4.5) | 0 | 0 | (0; 13.2) | 0 | 0 | 0 (0; 4.2) | 0 | 0 | 0 (0; 18.5) | 0 | 0 | 0 (0; 11.9) | 0 |
| AESI | 0 | 0 (0; 13.2) | 0 | 0 | 0 (0; 4.5) | 0 | 0 | (0; 13.2) | 0 | 0 | 0 (0; 4.2) | 0 | 0 | 0 (0; 18.5) | 0 | 0 | 0 (0; 11.9) | 0 |
| Grade 3 | 0 | 0 (0; 13.2) | 0 | 0 | 0 (0; 4.5) | 0 | 0 | (0; 13.2) | 0 | 0 | 0 (0; 4.2) | 0 | 0 | 0 (0; 18.5) | 0 | 0 | 0 (0; 11.9) | 0 |
| MAAE | 1 | 3.8 (0.1; 19.6) | 1 | 5 | 6.3 (2.1; 14.0) | 8 | 2 | (0.9; 25.1) | 3 | 3 | 3.5 (0.7; 10.0) | 5 | 0 | 0 (0; 18.5) | 0 | 1 | 3.4 (0.1; 17.8) | 1 |
| Grade 3 | 0 | 0 (0; 13.2) | 0 | 1 | 1.3 (0; 6.8) | 1 | 0 | (0; 13.2) | 0 | 0 | (0; 4.2) | 0 | 0 | 0 (0; 18.5) | 0 | 0 | 0 (0; 11.9) | 0 |

#### B. Single-dose cohort

| | LD + AF03 N=36 | | | LD + AS03 N=36 | | | HD + AF03 N=35 | | | HD + AS03 N=34 | | | Placebo N=34 | | |
|---|---|---|---|---|---|---|---|---|---|---|---|---|---|---|---|
| | **n** | **% (95% CI)** | **AEs** | **n** | **% (95% CI)** | **AEs** | **n** | **% (95% CI)** | **AEs** | **n** | **% (95% CI)** | **AEs** | **n** | **% (95% CI)** | **AEs** |
| Unsolicited AE | 10 | 27.8 (14.2; 45.2) | 15 | 9 | 25.0 (12.1; 42.2) | 17 | 9 | 25.7 (12.5; 43.3) | 18 | 10 | 29.4 (15.1; 47.5) | 14 | 8 | 23.5 (10.7; 41.2) | 12 |
| Grade 3 | 0 | 0 (0; 9.7) | 0 | 1 | 2.8 (0.1; 14.5) | 1 | 0 | 0 (0; 10.0) | 0 | 0 | 0 (0; 10.3) | 0 | 0 | 0 (0; 10.3) | 0 |
| Unsolicited AR | 5 | 13.9 (4.7; 29.5) | 6 | 3 | 8.3 (1.8; 22.5) | 3 | 4 | 11.4 (3.2; 26.7) | 7 | 3 | 8.8 (1.9; 23.7) | 3 | 1 | 2.9 (0.1; 15.3) | 1 |
| Grade 3 | 0 | 0 (0; 9.7) | 0 | 0 | 0 (0; 9.7) | 0 | 0 | 0 (0; 10.0) | 0 | 0 | 0 (0; 10.3) | 0 | 0 | 0 (0; 10.3) | 0 |
| Unsolicited injection site AR | 2 | 5.6 (0.7; 18.7) | 3 | 0 | 0 (0; 9.7) | 0 | 1 | 2.9 (0.1; 14.9) | 1 | 2 | 5.9 (0.7; 19.7) | 2 | 0 | 0 (0; 10.3) | 0 |
| Grade 3 | 0 | 0 (0; 9.7) | 0 | 0 | 0 (0; 9.7) | 0 | 0 | 0 (0; 10.0) | 0 | 0 | 0 (0; 10.3) | 0 | 0 | 0 (0; 10.3) | 0 |
| Unsolicited systemic AE | 8 | 22.2 (10.1; 39.2) | 12 | 9 | 25.0 (12.1; 42.2) | 17 | 9 | 25.7 (12.5; 43.3) | 17 | 9 | 26.5 (12.9; 44.4) | 12 | 8 | 23.5 (10.7; 41.2) | 12 |
| Grade 3 | 0 | 0 (0; 9.7) | 0 | 1 | 2.8 (0.1; 14.5) | 1 | 0 | 0 (0; 10.0) | 0 | 0 | 0 (0; 10.3) | 0 | 0 | 0 (0; 10.3) | 0 |
| Unsolicited systemic AR | 3 | 8.3 (1.8; 22.5) | 3 | 3 | 8.3 (1.8; 22.5) | 3 | 3 | 8.6 (1.8; 23.1) | 6 | 1 | 2.9 (0.1; 15.3) | 1 | 1 | 2.9 (0.1; 15.3) | 1 |
| Grade 3 | 0 | 0 (0; 9.7) | 0 | 0 | 0 (0; 9.7) | 0 | 0 | 0 (0; 10.0) | 0 | 0 | 0 (0; 10.3) | 0 | 0 | 0 (0; 10.3) | 0 |
| SAE | 1 | 2.8 (0.1; 14.5) | 1 | 0 | 0 (0; 9.7) | 0 | 1 | 2.9 (0.1; 14.9) | 1 | 0 | 0 (0; 10.3) | 0 | 0 | 0 (0; 10.3) | 0 |
| Grade 3 | 0 | 0 (0; 9.7) | 0 | 0 | 0 (0; 9.7) | 0 | 0 | 0 (0; 10.0) | 0 | 0 | 0 (0; 10.3) | 0 | 0 | 0 (0; 10.3) | 0 |
| AESI | 0 | 0 (0; 9.7) | 0 | 1 | 2.8 (0.1; 14.5) | 1 | 0 | 0 (0; 10.0) | 0 | 0 | 0 (0; 10.3) | 0 | 0 | 0 (0; 10.3) | 0 |
| Grade 3 | 0 | 0 (0; 9.7) | 0 | 0 | 0 (0; 9.7) | 0 | 0 | 0 (0; 10.0) | 0 | 0 | 0 (0; 10.3) | 0 | 0 | 0 (0; 10.3) | 0 |
| MAAE | 2 | 5.6 (0.7; 18.7) | 3 | 3 | 8.3 (1.8; 22.5) | 4 | 1 | 2.9 (0.1; 14.9) | 1 | 1 | 2.9 (0.1; 15.3) | 1 | 3 | 8.8 (1.9; 23.7) | 3 |
| Grade 3 | 0 | 0 (0; 9.7) | 0 | 0 | 0 (0; 9.7) | 0 | 0 | 0 (0; 10.0) | 0 | 0 | 0 (0; 10.3) | 0 | 0 | 0 (0; 10.3) | 0 |

Numbers presented are numbers of participants experiencing at least one of the specified items, or the number of adverse events specified (column, AEs).
AESI, adverse event of special interest; AR, adverse reaction; HD, high-dose; LD, high-dose; MAAE, medically attended adverse event; (S)AE, serious adverse event

**Supplementary Figure S1: Study design for (A) the single-dose and (B) the two-dose schedule**

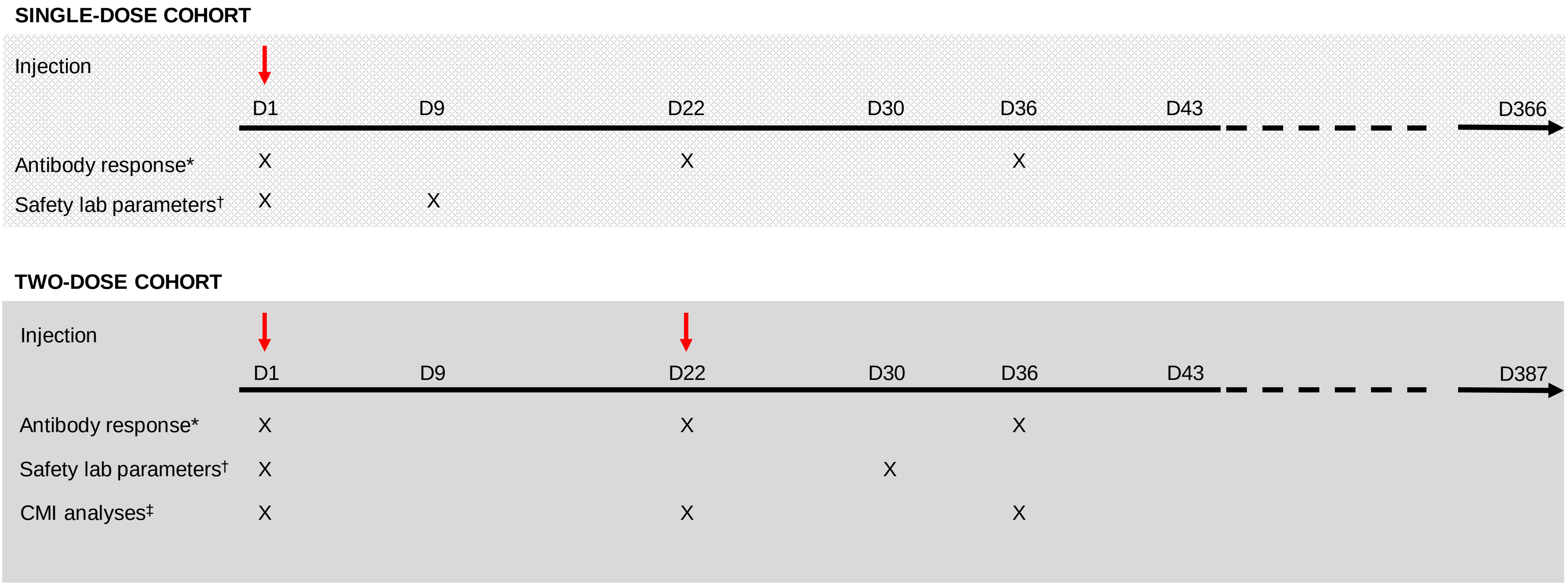


CMI, cell-mediated immunity; D, study day; red arrows indicate the first or second study injections.

*Serum samples (30 mL) were used to determine SARS-CoV-2 neutralising and binding antibody responses at the specified timepoints; [†]Assessments included serum chemistries, haematology, urinalysis and microscopy; [‡]Whole blood samples (10 mL) from a subset of participants in the 18–49 years age stratum in the two-dose cohort were used for cell-mediated immunity analyses.

**Supplementary Figure S2: Participant flow through the study for those randomised to receive a single dose in the (A) 18-49 years and (B) ≥ 50 years age strata.**

A

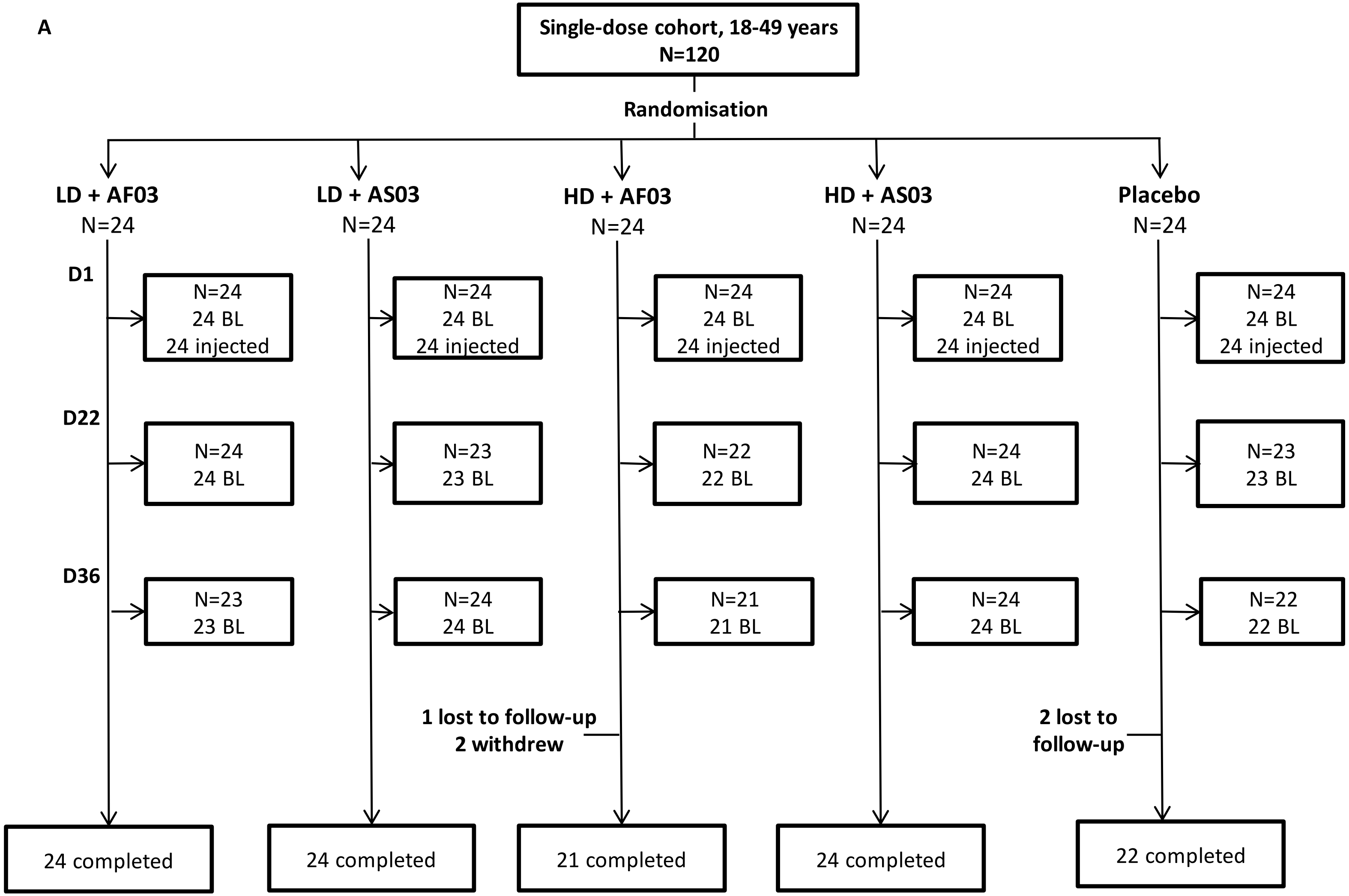

B

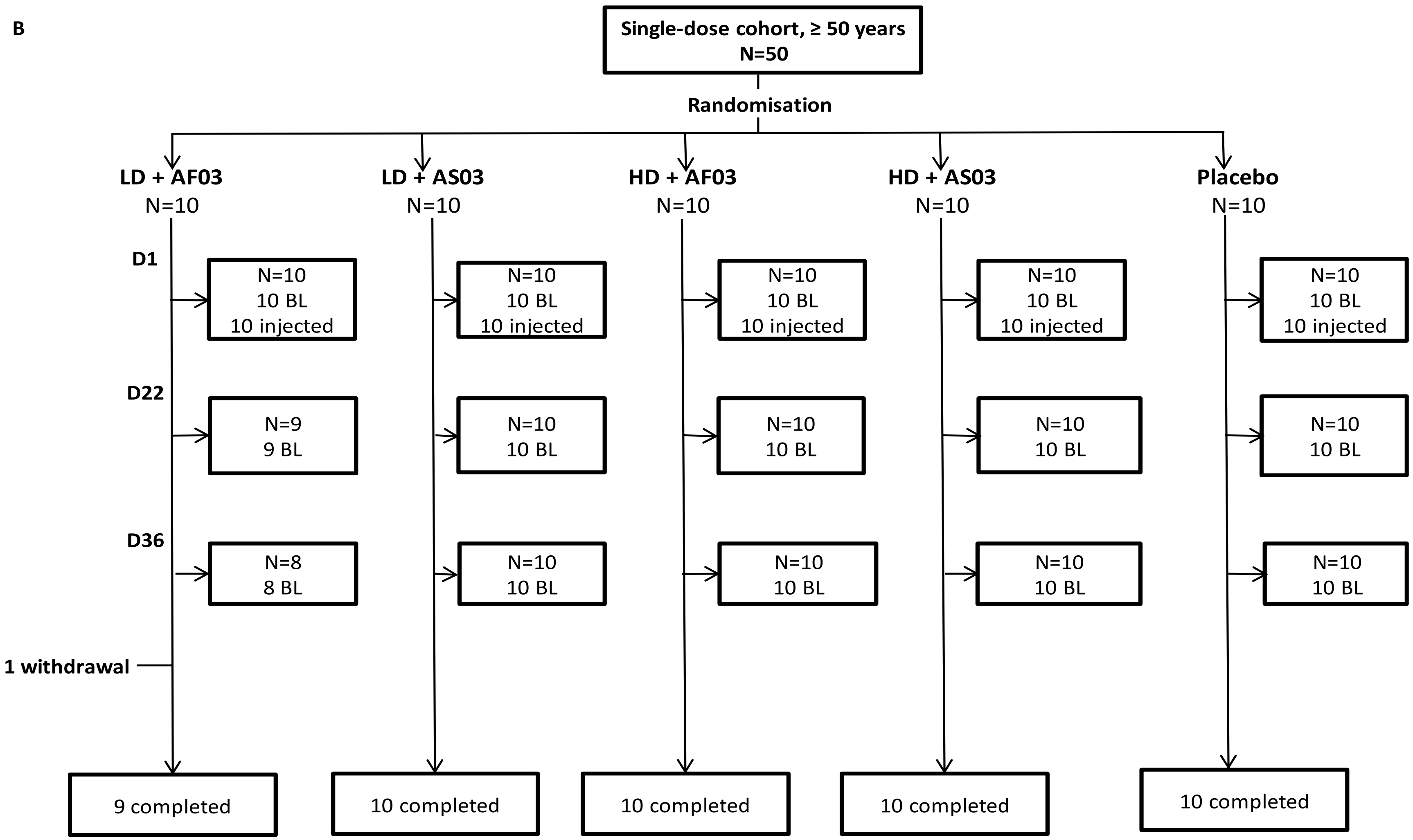
Single-dose cohort, ≥ 50 years
N=50
Randomisation
LD + AF03
N=10
LD + AS03
N=10
HD + AF03
N=10
HD + AS03
N=10
Placebo
N=10
D1
N=10
10 BL
10 injected
N=10
10 BL
10 injected
N=10
10 BL
10 injected
N=10
10 BL
10 injected
N=10
10 BL
10 injected
D22
N=9
9 BL
N=10
10 BL
N=10
10 BL
N=10
10 BL
N=10
10 BL
D36
N=8
8 BL
N=10
10 BL
N=10
10 BL
N=10
10 BL
N=10
10 BL
1 withdrawal
9 completed
10 completed
10 completed
10 completed
10 completed

**Supplementary Figure S3. Solicited injection site reactions (A) and solicited systemic reactions (B) after a single dose, by age strata (single-dose cohort, SafAS)**

**A.**

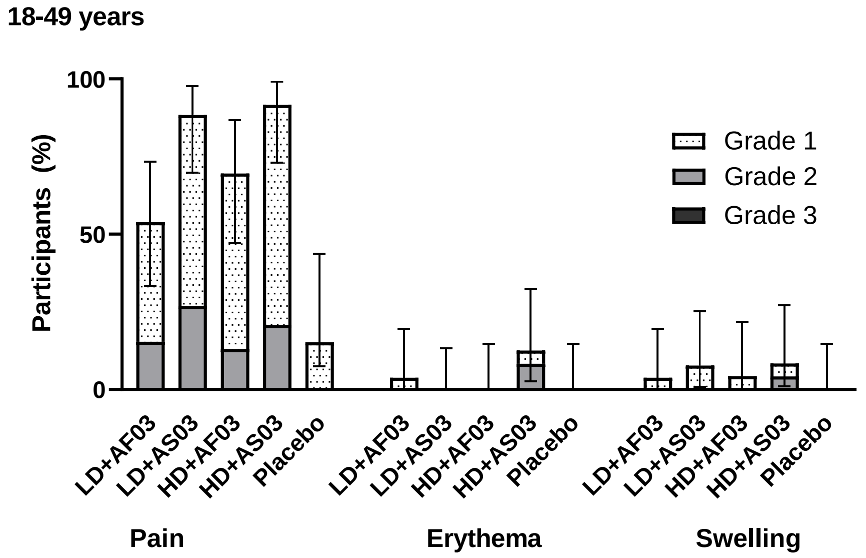


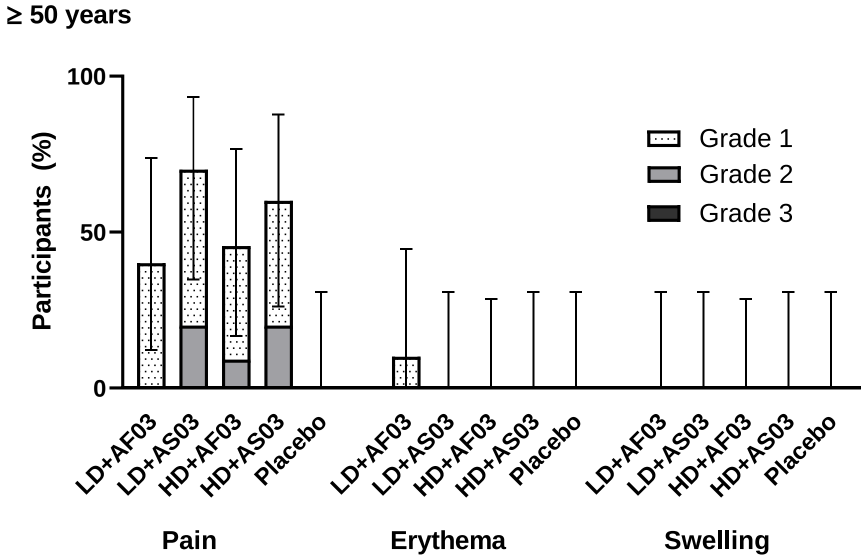

**B.**

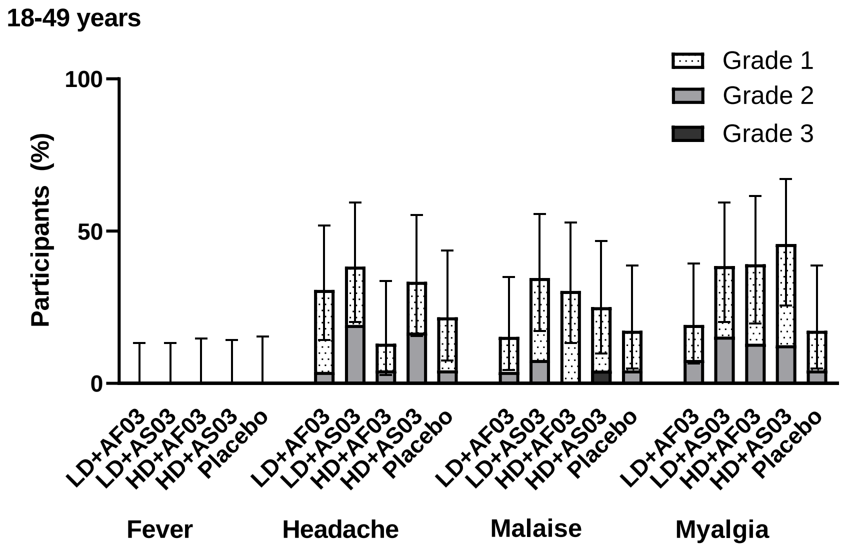
18-49 years
Grade 1
Grade 2
Grade 3
Participants (%)
100
50
0
LD+AF03
LD+AS03
HD+AF03
HD+AS03
Placebo
Fever
Headache
Malaise
Myalgia

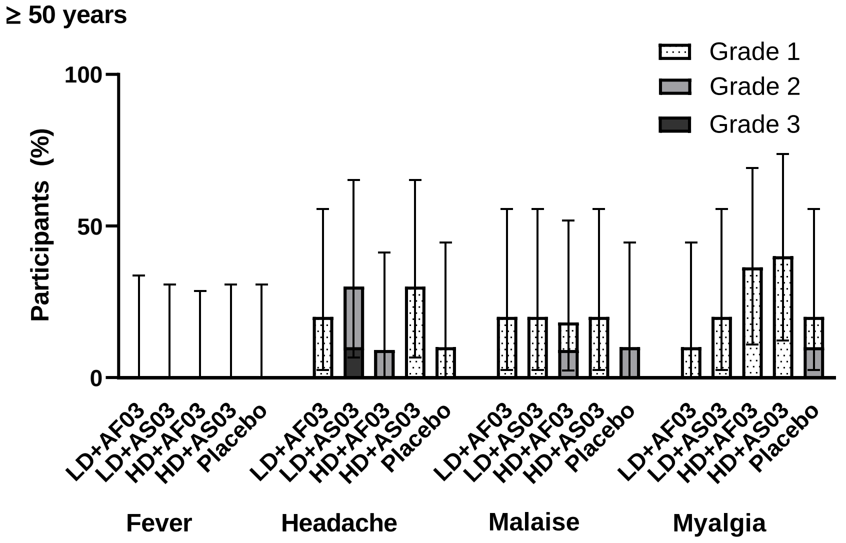
≥ 50 years
Grade 1
Grade 2
Grade 3
Participants (%)
100
50
0
LD+AF03
LD+AS03
HD+AF03
HD+AS03
Placebo
Fever
Headache
Malaise
Myalgia

**Supplementary Figure S4. Neutralising antibody titres (microneutralisation assay; A) and binding antibody responses (ELISA; B) after single dose, by age strata (single-dose cohort; PPAS-IAS)**

Footnote: Neutralising antibody titres are additionally shown for a panel of 93 convalescent plasma samples (A).

**A**

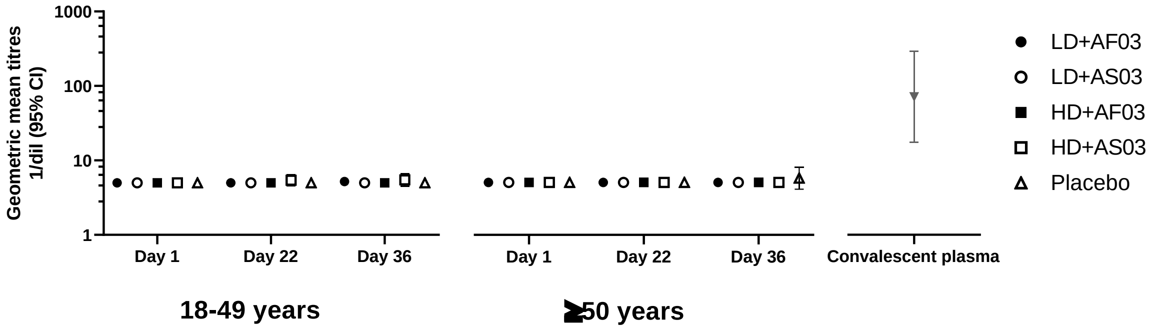


**B**

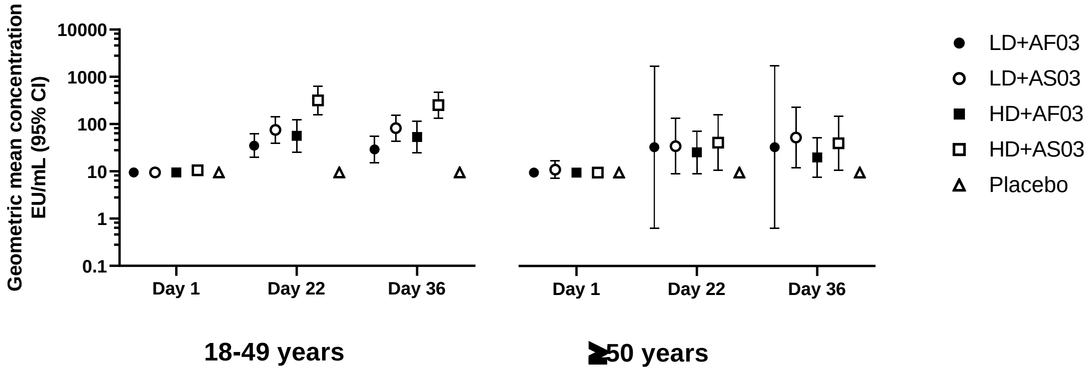

**Supplementary Figure S5. Percentages of participants with (A) a 4-fold rise in neutralising antibody titres, relative to D1, and (B) seroconversion, overall and by age strata at Day 22 and Day 36, two dose cohort (PPAS-IAS)**

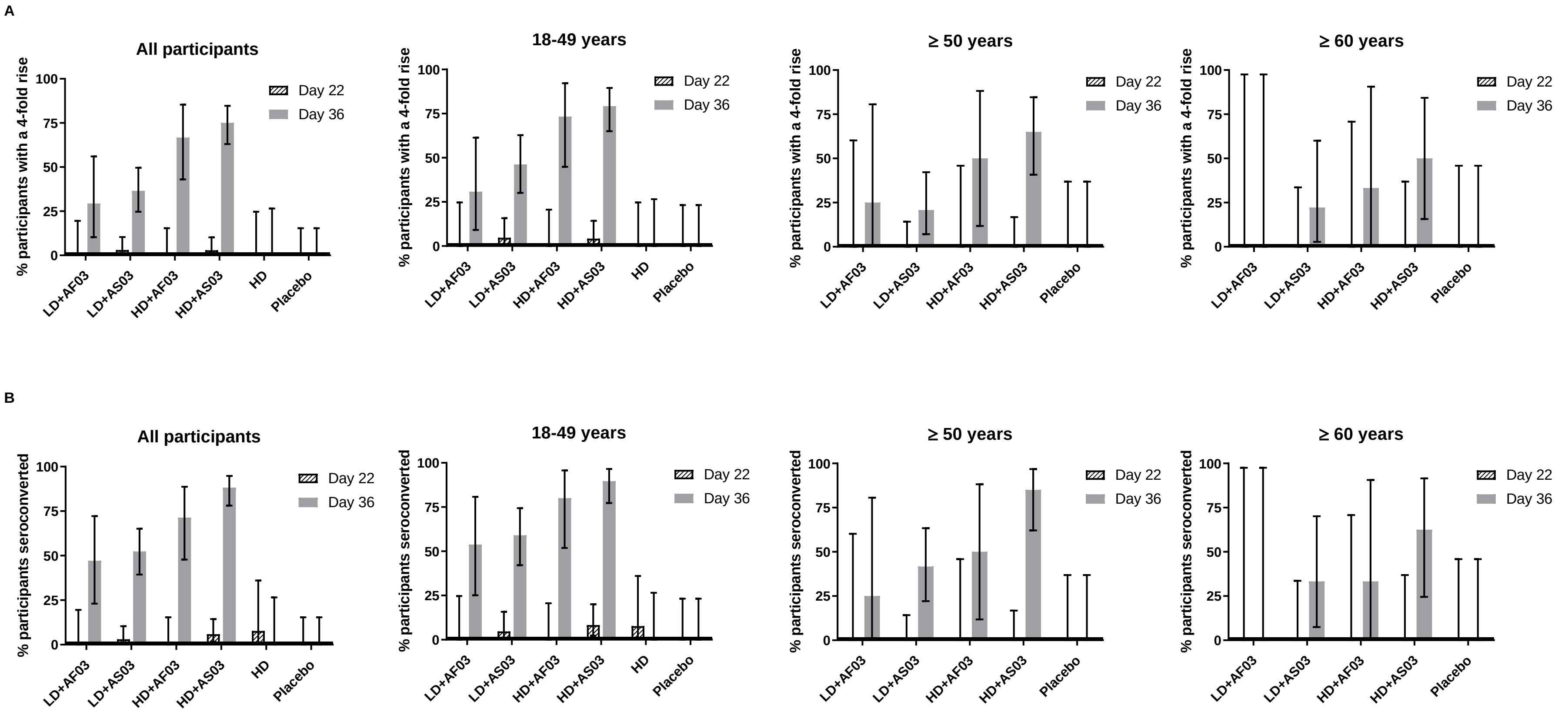

**Supplementary Figure S6. CMI analyses – ratios of fold-rises for IFN-γ (A), IL-2 (B) and TNFα (C) to IL-4, IL-5 and IL-13 at Day 22 and Day 36 (PPAS-CMI)**

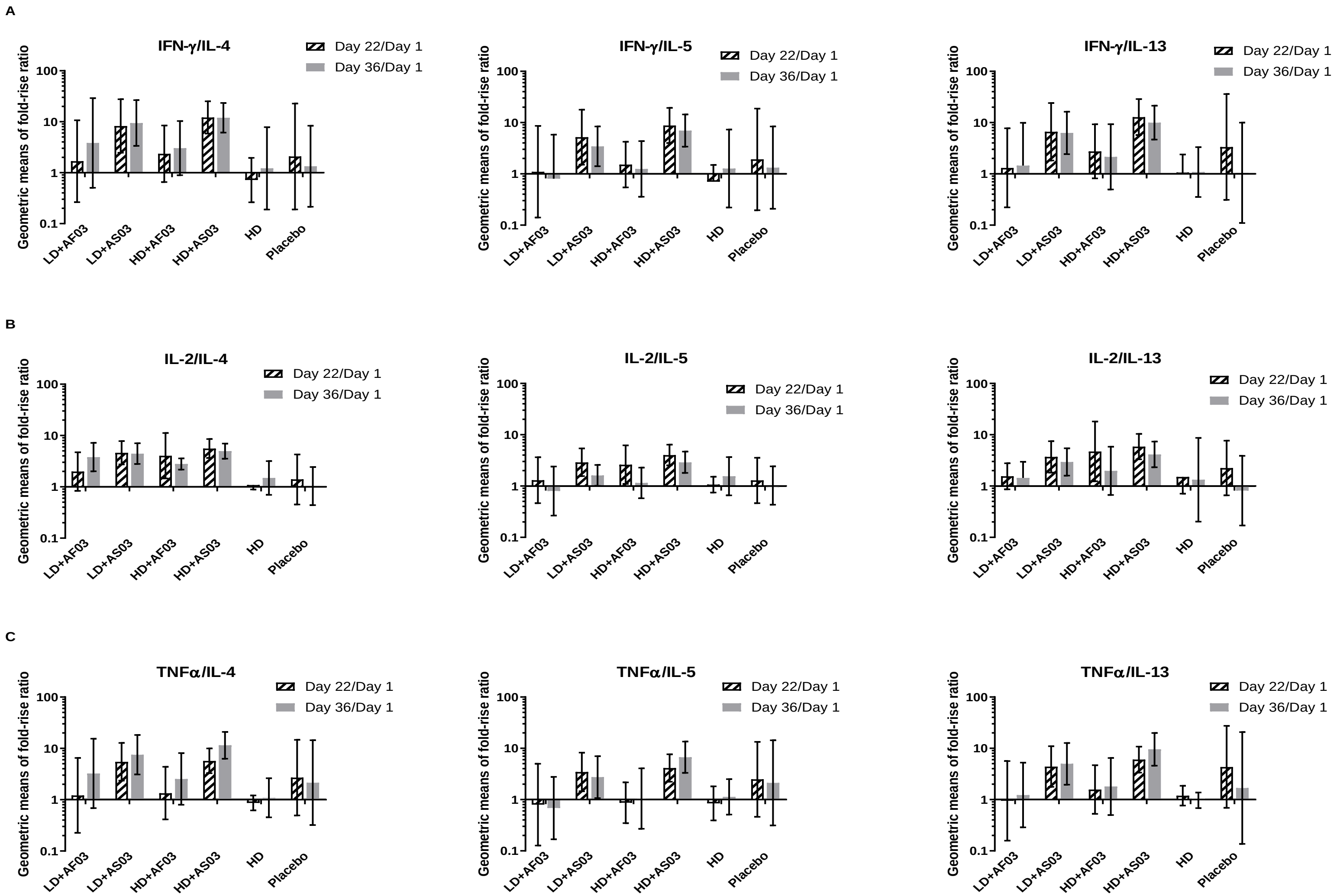
